# 2-Deoxy-D-Glucose as an Adjunct to Standard of Care in the Medical Management of COVID-19: A Proof-of-Concept & Dose-Ranging Randomised Clinical Trial

**DOI:** 10.1101/2021.10.08.21258621

**Authors:** Anant Narayan Bhatt, Srinivas Shenoy, Sagar Munjal, Vijayakumar Chinnadurai, Apurva Agarwal, A. Vinoth Kumar, A Shanavas, Ratnesh Kanwar, Sudhir Chandna

**Author notes:** **Corresponding Authors:** 1. Anant Narayan Bhatt, Institute of Nuclear Medicine and Allied Sciences, Defence Research and Development Organization, Timarpur, Delhi – 110054, India,;, 2. Srinivas Shenoy, Dr Reddy’s Laboratories Limited, 8-2-337, Road No.3, Banjara Hills, Hyderabad-500 034, India.

## Abstract

**Objective:** To evaluate the efficacy and safety of 2-deoxy-D-glucose (2-DG) in the treatment of COVID-19.

**Participants:** 110 adults aged 18 to 65 years with moderate to severe COVID-19.

**Interventions:** 63, 90, and 126 mg/kg/day 2-DG plus standard of care (SOC) versus SOC only.

**Main Outcome Measures:** Times to maintaining SpO_2_ ≥94% on room air discharge, clinical recovery, vital signs normalisation, improvement by 1 and 2 points on WHO 10-point ordinal scale, negative conversion on RT-PCR, intensive care, and mortality.

**Results:** Patients treated with 90 mg/kg/day 2-DG plus SOC showed better outcomes. Time to maintaining SpO2 ≥94% was significantly shorter in the 2-DG 90 mg compared to SOC (median 2.5 days vs 5 days, Hazard ratio [95% confidence interval]=2.3 [1.14, 4.64], *p*=0.0201). Times to discharge from isolation ward, to clinical recovery, and to vital signs normalisation were significantly shorter for the 2-DG 90 mg group.

All three doses of 2-DG were well tolerated. Thirty-three (30.3%) patients reported 65 adverse events and were mostly (86%) mild.

**Conclusion:** 2-DG 90 mg/kg/day as adjunct to SOC showed clinical benefits over SOC alone in the treatment of moderate to severe COVID-19. The promising trends observed in current phase-II study encourage confirmatory evaluation of the efficacy and safety of 2-DG in a larger phase-III trial.

## INTRODUCTION

Coronavirus disease 2019 (COVID-19) is currently a major global public health crisis. While remarkable progress has been made in vaccine development, there are limited therapeutic interventions available. Although several treatment modalities have been tried, no curative treatment has been found to date for COVID-19, and it is increasingly apparent that a multimodal approach is necessary for acute COVID-19 management [1].

The synthetic glucose analogue 2-deoxy-D-glucose (2-DG) has been identified as a potential treatment for COVID-19. It inhibits glycolysis in host cells infected by the severe acute respiratory syndrome coronavirus 2 (SARS-CoV-2) [2 - 4].

Upon infecting host cells, viruses reprogram host cell metabolism for their own rapid replication [5]. New virion assembly requires high levels and turnover of nucleotides and lipids, which are achieved by elevated levels of glucose transporters and enhanced aerobic glycolysis (i.e., Warburg effect) [6]. Administration of 2-DG leads to its preferential accumulation within virally infected host cells, on account of the higher number of glucose transporters on the membranes of these virally infected cells, as compared to uninfected cells. Subsequently, 2-DG blocks glycolysis, resulting in the depletion of adenosine triphosphate (ATP) and anabolic intermediates required for viral replication and packaging. In addition to this direct effect, 2-DG may also cause mis-glycosylation of nascent viral proteins to form defective progeny virions with low potential to secondarily infect neighbouring cells [2, 7]. Moreover, 2-DG also exerts anti-inflammatory effects and was shown to inhibit viral infection and inflammation in lungs in a murine model [8].

The antiviral activity of 2-DG was demonstrated in a study in 36 women with herpes simplex infection, when applied topically [9]. Furthermore, in vitro studies have shown significant inhibition of SARS-CoV-2 replication by 2-DG [2 - 4].

Recent studies using Positron Emission Tomography (PET) with the radiotracer, ^18^FDG (fluorodeoxyglucose, an analogue of 2-DG) demonstrates substantial accumulation of the radiolabeled FDG in the inflamed lungs of COVID-19 patients [10,11]. This indicates that 2-DG could also preferentially and disproportionately accumulate in the inflamed lungs of COVID-19 patients. Furthermore, 2-DG has been studied in several clinical trials for treatment of various cancers globally and has demonstrated acceptable tolerability in humans [12].

This phase-II clinical trial was conducted to evaluate the efficacy and safety of 2-DG as adjunctive therapy in the treatment of moderate to severe COVID-19. The starting daily dose of 2-DG, 63 mg/kg was chosen based on tolerability data from previous clinical studies in patients with solid tumours [13,14]. Dose escalation to 90 mg/kg/day (nearly 1.5x) and 126 mg/kg/day (2x) were planned if no safety concerns were observed at the starting dose. It should be noted that tolerability up to 250 mg/kg was established in a previous clinical study in glioblastoma multiforme [15].

## METHODS

This was a phase-II, multicentre, randomised, open-label, parallel-group clinical trial to evaluate the efficacy, safety, and tolerability of 2-DG administered adjunctly to standard of care (SOC), in comparison with SOC alone, in patients with moderate or severe COVID-19. SOC was based on the national guideline [16].

The trial was conducted under the supervision of Drugs Controller General of India (DCGI) and approved by appropriate ethics committees (EC). The sample size (110 patients, 22 subjects in each arm) for this proof-of-concept study was mutually decided upon between the sponsor and DCGI based on the novelty of the drug and with limited efficacy and safety information available in non-cancer patients. The trial was prospectively registered on Clinical Trials Registry – India [CTRI/2020/06/025664 (Registered on: 05/06/2020)].

### Participating Patients

The trial enrolled male or female patients aged 18 to 65 years, who were admitted to isolation wards at 12 COVID-19 management hospitals in India. Critically ill patients as defined in the guidance were excluded from the study [16]. The diagnosis of COVID-19 was confirmed by real-time reverse transcriptase polymerase chain reaction (rRT-PCR) assay of each patient’s nasopharyngeal/oropharyngeal swab specimen.

COVID-19 severity in each patient was assessed according to the guidance [16]. Moderate disease was defined as the presence of dyspnoea and/or hypoxia, fever, cough, including an oxygen saturation level (SpO_2_) of 90% to 94% on room air, and a respiratory rate of ≥ 24 per minute. Severe disease was defined as the presence of clinical signs of pneumonia plus one of the following: respiratory rate > 30 per minute, severe respiratory distress, SpO_2_ < 90% on room air, but not critically ill, i.e., no acute respiratory distress syndrome (ARDS), multiorgan failure, or septic shock. Since, 2-DG has previously been reported to cause QT prolongation, patients with cardiac conduction delay (QT_c_ > 500 msec) or patients taking any medications known to prolong QT interval (including hydroxychloroquine and azithromycin), were not included in the study. Patients with malabsorption/gastrointestinal abnormalities that may affect drug absorption, and patients with body weight < 45 kg or > 130 kg were excluded from the study.

### Trial Design

The trial was conducted in two parts, Part A for proof-of-concept (clinical), and Part B for dose ranging. Powder form of 2-DG was dissolved in 100 mL of potable water and an individualised volume of the solution was dosed orally to patients based on body weight. In Part A, 44 patients were randomised in a 1:1 ratio to receive treatment with either 2-DG 63 mg/kg/day plus SoC (the 2-DG 63 mg group) or SOC only (the SOC1 group). The centralized randomisation was carried out manually using Statistical Analysis Software (SAS, version 9.4), throughout the study. 2-DG was given in two split doses totaling 63 mg/kg/day, viz., 45 mg/kg in the morning and 18 mg/kg in the evening, along with SOC, until the patient was discharged or up to 28 days after the initiation of study treatment (i.e., Day 1), whichever occurred first. In the SOC1 group, SOC was provided as long as required. The dose-ranging Part B was conducted after the safety results from Part A had been reviewed and deemed acceptable. A total of 66 patients were randomised in a 1:1:1 ratio to receive 2-DG 90 mg/kg/day plus SOC (the 2-DG 90 mg group), 2-DG 126 mg/kg/day plus SOC (the 2-DG 126 mg group), and SOC only (SOC2 group). 2-DG was administered in two equally divided doses in the morning and evening, viz., 45mg/kg for 90mg group, and 63 mg/kg for 126mg group. In the two active treatment groups, 2-DG was administered along with SOC for 10 days or until discharge, whichever occurred earlier. In the SOC2 group, SOC was provided as long as required.

For both parts of the study, data were collected through 28 days of study or until a patient was discharged from isolation ward of the hospital, whichever occurred earlier. A patient’s clinical status was evaluated every day by the treating physician using the World Health Organization (WHO) 10-point ordinal scale [17]. The following assessments were done daily; the severity of COVID-19-associated symptoms, vital signs, peripheral blood oxygen saturation (SpO_2_), partial physical examination, 12-lead ECG, random blood glucose, adverse events, and concomitant medication. Real-time RT-PCR assay was carried out on nasopharyngeal/oropharyngeal swab samples on Days 3, 7, 10, 14, and 28 (or day of discharge if earlier) during Part A of the study and on Days 1, 3, 5, 7, 9, 10, 14, and 28 (or day of discharge if earlier) during Part B. Clinical laboratory tests (haematology, serum biochemistry, and urinalysis) were performed on Days 7, 14, and 28 (or discharge if earlier). The severity of COVID-19-associated symptoms of cough, fever, nasal congestion, fatigue, body aches, sore throat, breathlessness, and diarrhea were self-scored by the patient every day using a 5-point Likert-type scale (0= absent, 1= mild, 2=moderate, 3= severe,4= very severe/critical) for each symptom.

A Data Safety Monitoring Board (DSMB) was instituted to review the safety data throughout the trial, as per the DSMB charter.

### Statistical Methods

Continuous data were summarised using descriptive statistics. Categorical data were summarised using counts and percentage. ‘Time-to-event’ analyses compared between treatment and control (2-DG plus SOC versus SOC) groups using the Cox proportional hazard (CPH) model, with baseline clinical status scores as covariates. Age and sex were considered as additional covariates wherever relevant. Median estimates, hazard ratio (HR) and its corresponding two-sided 95% confidence interval (CI), and two-sided *p* values at 5% level of significance are reported. Statistical comparisons were done using log-rank test and Kaplan-Meier plot wherever applicable. Proportions were compared using Chi-square or Fisher exact test.

There were no statistical power calculations for sample size. Efficacy data were collected based on several clinically meaningful measures, and no particular parameter was designated as the primary endpoint.

The 2-DG plus SOC treatment groups were compared with their contemporaneous SOC groups as well as with the pooled SOC (SOC1 + SOC2).

## RESULTS

A total of 110 patients were randomised, between June 2019 and September 2020, with 22 patients in each of the five treatment groups: the 2-DG 63 mg and SOC1 groups in Part A and the 2-DG 90 mg, 2-DG 126 mg, and SOC2 group in Part B. 109 were dosed, and 1 patient in the 2-DG 126 mg group was discontinued before receiving any dose, due to an adverse event (Figure 1).

**Figure 1:**
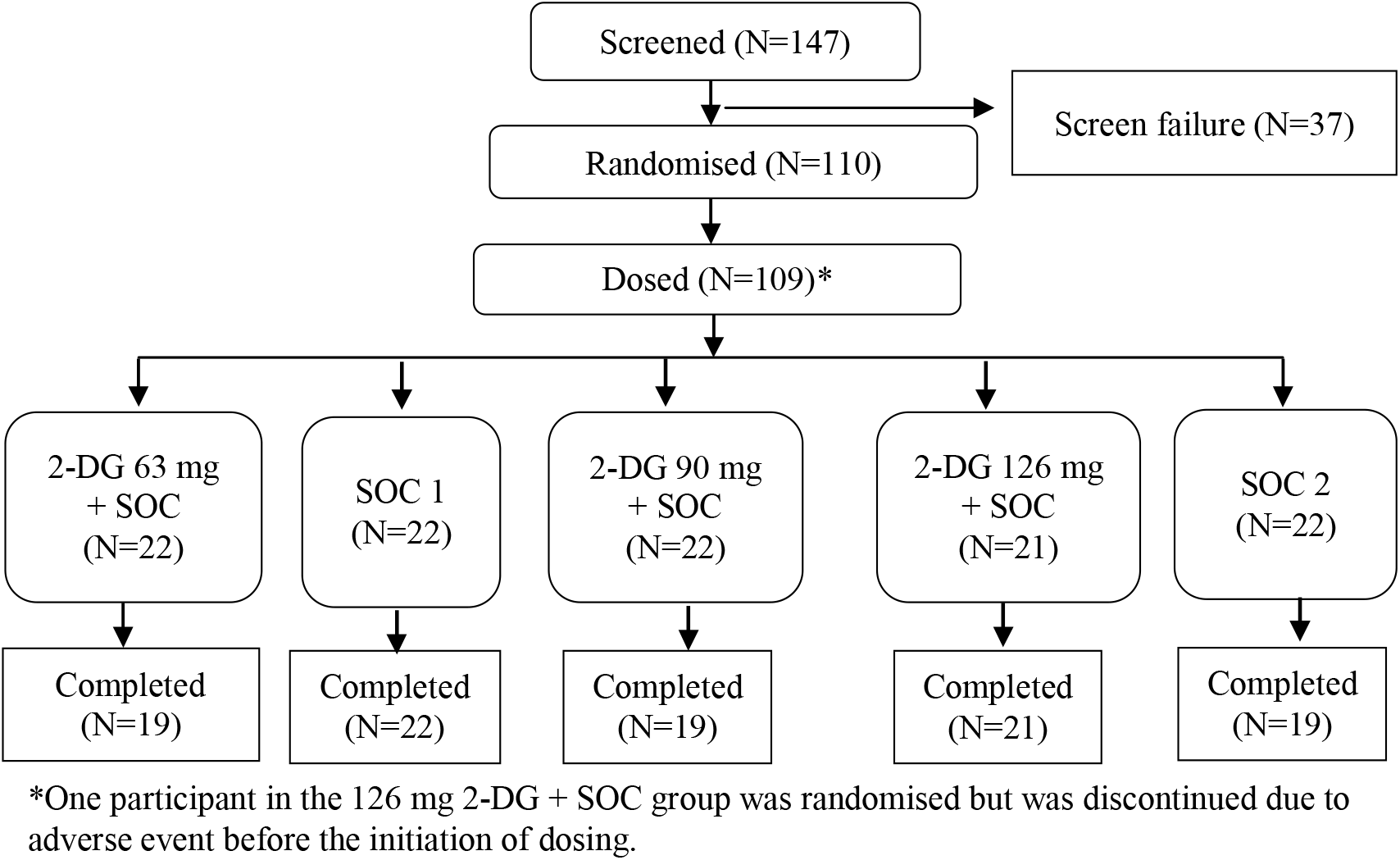
Study flow chart displaying patient counts in each treatment group. Abbreviations: 2-DG=2-deoxy-D-glucose, SOC=standard of care, SOC1=SOC in Part A of the study

The demographic and baseline characteristics were similar across the treatment groups (Table 1). The mean (standard deviation [SD]) age was 44.9 (10.90) years, and the mean (SD) weight was 68.6 (11.39) kg for all randomised patients. A majority of the patients (88 [80.7%]) were male. There were differences in certain baseline disease characteristics between patients enrolled in Parts A and B of the study. The mean (SD) number of days since the onset of initial COVID-19 symptoms in patients enrolled in Part A were 7.2 (2.58) days for the SOC1 group and 6.6 (2.26) days for the 2-DG 63 mg group, and for those enrolled in Part B were 4.5 (1.41) days, 4.3 (1.46) days, and 4.4 (1.40) days for the 2-DG 90 mg, 2-DG 126 mg, and SOC2 groups, respectively. All enrolled patients were assessed to have moderate COVID-19 as defined by the guidance, except 3 patients with missing severity data [one (4.5%) patient each from 2-DG 63 mg, SOC1, and SOC2 groups].

**Table 1:** Patient Demographic and Baseline Characteristics.

|  |  | **2-DG 63mg+ SOC** | **SOC1** | **2-DG 90mg SOC** | **2-DG 126mg SOC** | **SOC2** |
| --- | --- | --- | --- | --- | --- | --- |
|  |  | **N=22** | **N=22** | **+** | **+** | **N=22** |
|  |  | **n (%)** | **n (%)** | **N=22** | **N=21** | **n (%)** |
|  |  |  |  | **n (%)** | **n (%)** |  |
| Age (years) | Mean (SD) | 44.2 (12.71) | 44.4 (9.59) | 46.3 (11.00) | 42.7 (9.33) | 46.6 (11.96) |
| Weight (kg) | Mean (SD) | 61.3 (9.35) | 67.9 (9.76) | 69.4 (10.52) | 74.2 (12.23) | 70.6 (11.77) |
| Gender [n (%)] | Female | 7 (31.8) | 6 (27.3) | 1 (4.5) | 5 (23.8) | 2 (9.1) |
|  | Male | 15 (68.2) | 16 (72.7) | 21 (95.5) | 16 (76.2) | 20 (90.9) |
| Number of days since onset of first symptom of COVID-19 | Mean (SD) | 6.6 (2.26) | 7.2 (2.58) | 4.5 (1.41) | 4.3 (1.46) | 4.4 (1.40) |
| Clinical severity status as defined by MoH&FW [n (%)] | Group 1 (Mild) | 0 | 0 | 0 | 0 | 0 |
|  | Group 2 (Moderate) | 21 (95.5%)<sup>a</sup> | 21 (95.5%)<sup>a</sup> | 22 (100.0%) | 21 (100.0%) | 21 (95.5%)<sup>a</sup> |
|  | Group 3 (Severe) | 0 | 0 | 0 | 0 | 0 |
| Oxygen saturation (SpO<sub>2</sub>%) | N | 22 | 21 | 22 | 21 | 21 |
|  | Mean (SD) | 93.1 (2.39) | 93.0 (1.82) | 92.7 (1.55) | 92.5 (1.47) | 93.0(2.07) |
|  | Median | 92.0 | 93.0 | 93.0 | 93.0 | 93.0 |
|  |  | **N=22** | **N=22** | **+** | **+** | **N=22** |
|  |  | **n (%)** | **n (%)** | **N=22** | **N=21** | **n (%)** |
|  |  |  |  | **n (%)** | **n (%)** |  |
| Heart rate (beats per minute) | N | 22 | 21 | 22 | 21 | 21 |
|  | Mean (SD) | 85.0 (11.32) | 86.3 (15.17) | 81.3 (10.39) | 84.9 (11.57) | 89.6 (8.48) |
|  | Median | 80.0 | 84.0 | 80.0 | 88.0 | 89.0 |
| Respiratory rate (per minute) | N | 22 | 21 | 22 | 21 | 21 |
|  | Mean (SD) | 22.1 (3.04) | 21.4 (3.20) | 23.9 (2.41) | 24.4 (2.09) | 24.7± 2.61 |
|  | Median | 24.0 | 22.0 | 25.0 | 25.0 | 25.0 |
| WHO 10-point ordinal scale | N | 22 | 22 | 22 | 21 | 22 |
|  | Mean (SD) | 5.1 (0.35) | 5.0 (0.21) | 4.3 (0.48) | 4.3 (0.46) | 4.3 (0.48) |
|  | Median | 5.0 | 5.0 | 4.0 | 4.0 | 4.0 |

Abbreviations: 2-DG=2-deoxy-D-glucose, SOC=standard of care, SOC1=SOC in Part A of the study, SOC2=SOC in Part B of the study
N: Total number of patients in the specified treatment group.
n: Total number of patients in a given category.
Percentages are based on the total number of patients in the specified treatment group.
MoH&FW: Ministry of Health & Family Welfare, Government of India
<sup>a</sup>Severity assessment was missing for 1 patient each in these three groups.

### Efficacy

The median time to achieve and maintain SpO_2_ ≥ 94% on room air at sea level was the shortest in 2-DG 90 mg group (2.5 days), followed closely by the 2-DG 126 mg group (3.0 days). The median time to achieving SpO_2_ ≥ 94% was 5.0 days across all the other three groups, viz., 63 mg, SOC1 and SOC2. The hazard ratio (95% CI) for the 2-DG 90 mg group was 2.3 (1.14, 4.64) (*p*=0.0201) compared with the SOC2 group and 2.6 (1.49, 4.70) (*p*=0.0009) compared with the pooled SOC group. The comparisons between 2-DG 63 mg and SOC1 and between 2-DG 126 mg and SOC2 were not statistically significant.

The median time to discharge from isolation ward was 8.0 days in the 2-DG 90 mg group (Figure 2), which was the shortest among all groups (Table 2). The hazard ratio (95% CI) for the 2-DG 90 mg group was 2.2 (1.07, 4.70) (*p*=0.0336) compared with the SOC2 group and 2.2 (1.21, 4.04) (*p*=0.01) compared with the pooled SOC group. Other two dose groups (2-DG 63 mg and 2-DG 126 mg groups) were not statistically significant, when compared with their respective SOC groups.

**Figure 2:**
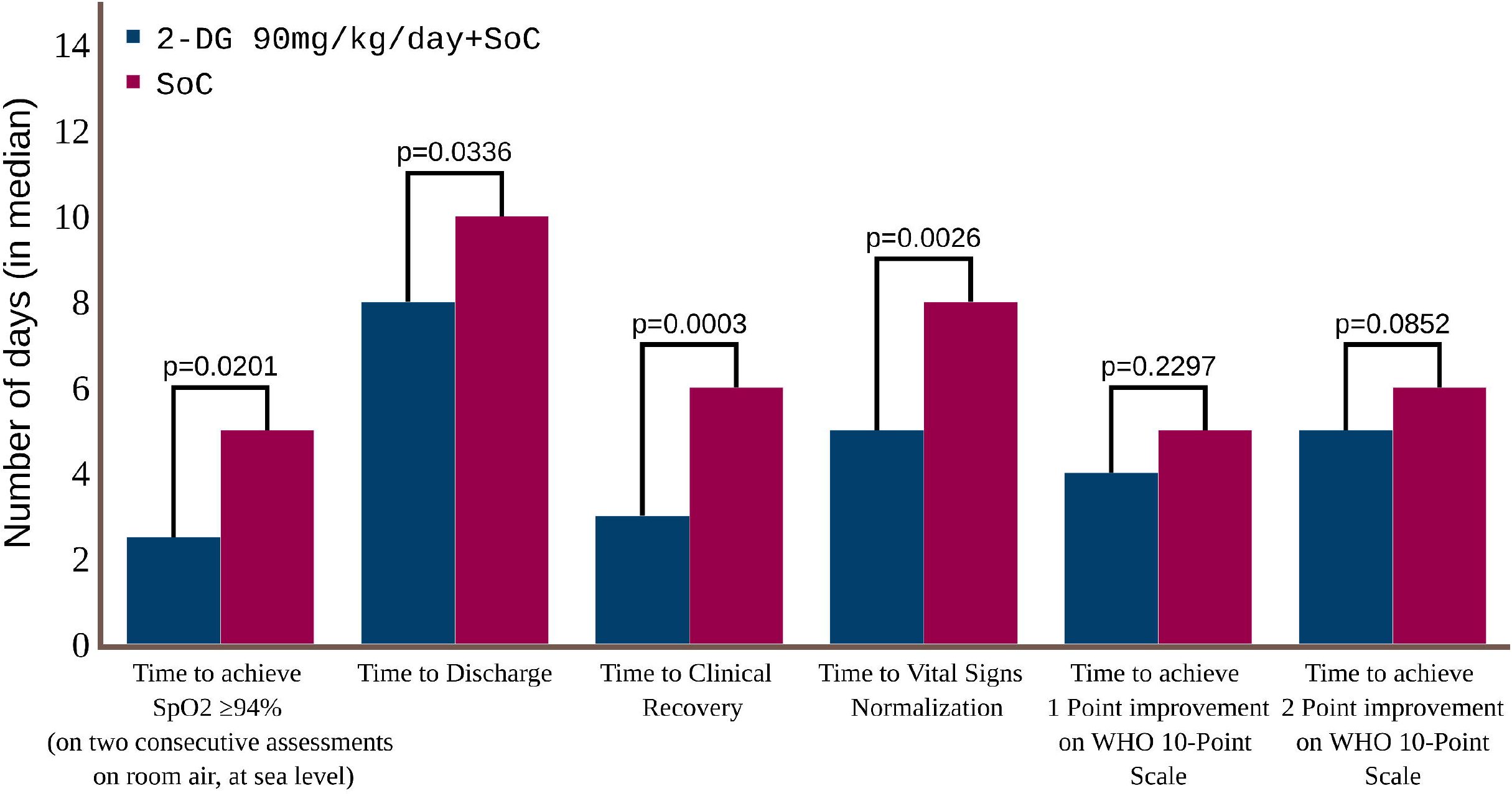
Median time (days) to clinical endpoints compared between patients receiving 2-DG 90 mg/kg/day plus SOC and patients receiving standard of care only. Abbreviations: 2-DG=2-deoxy-D-glucose, SOC=standard of care, WHO=World Health Organization, SOC2=SOC in Part B of the study

**Table 2:**
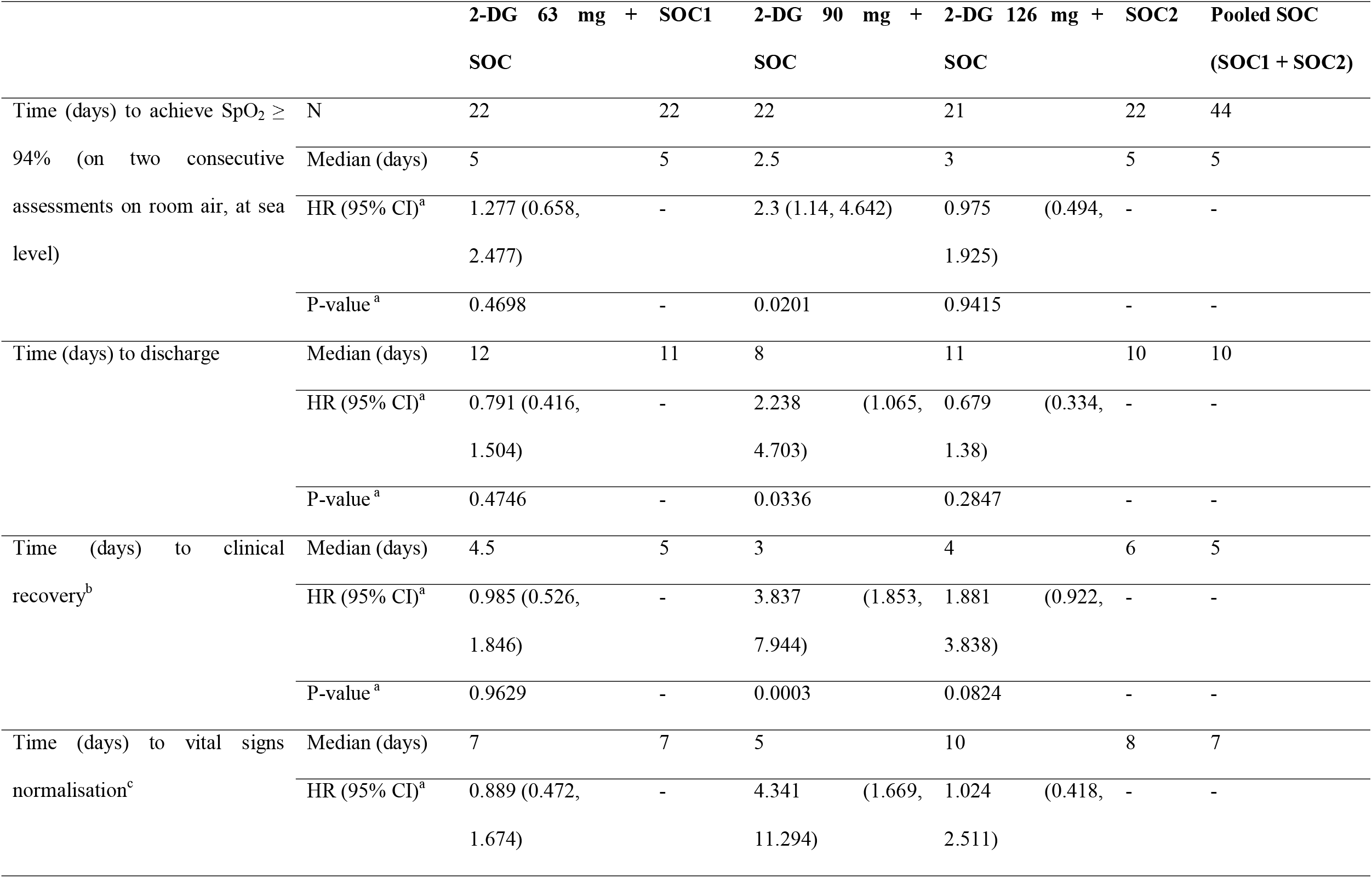
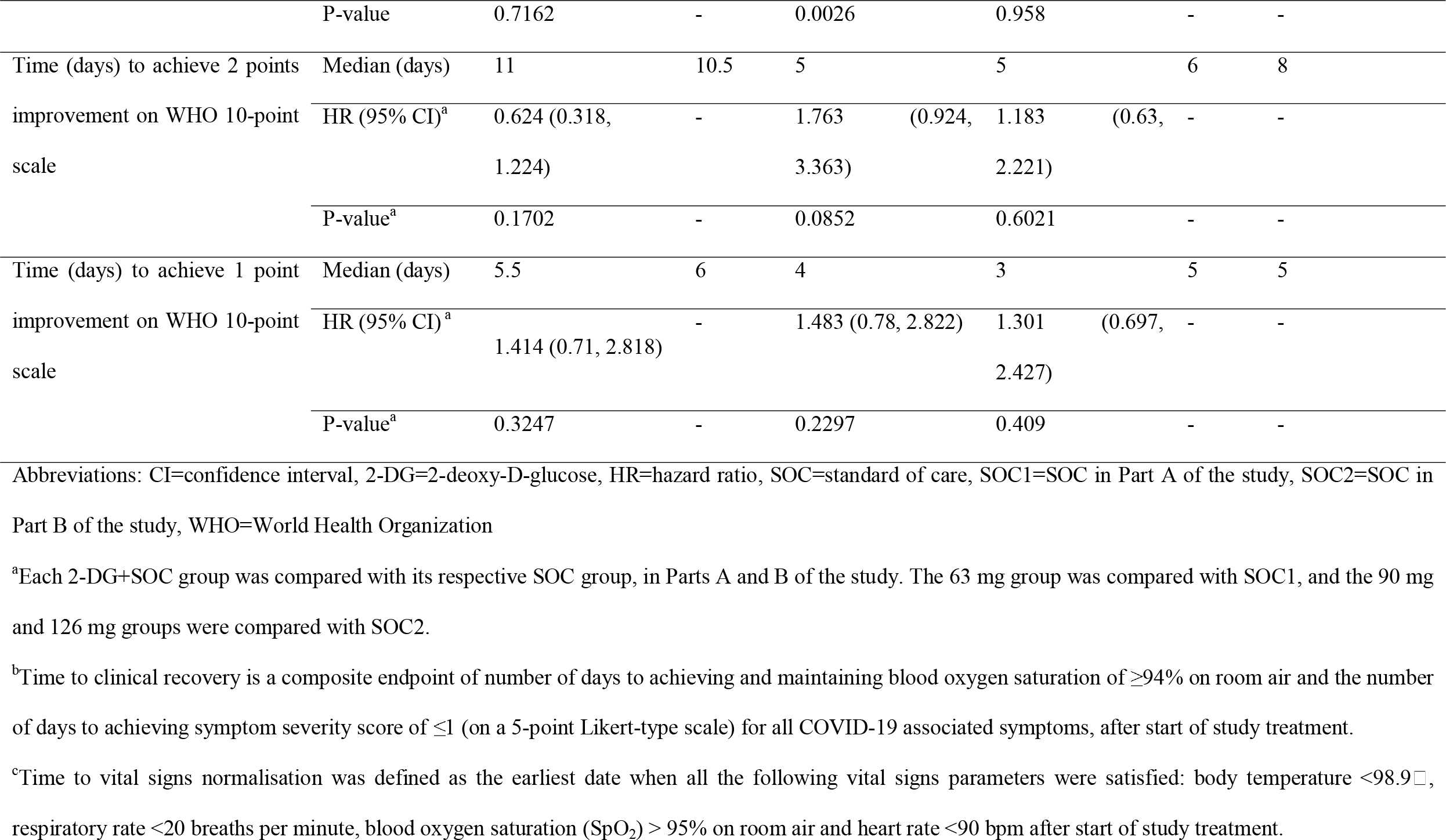
Efficacy endpoint comparisons between active (2-DG) and standard-of-care groups.

Time to clinical recovery is a composite endpoint of number of days to achieving and maintaining SpO_2_ of ≥ 94% on room air and the number of days to achieve symptom severity score (self-assessed by patient) of ≤1 on a 5-point Likert-type scale, for all COVID-19 associated symptoms.The median time to clinical recovery was 4.5 days, 3 days, and 4 days in the 2-DG 63 mg, 2-DG 90 mg, and 2-DG 126 mg groups, respectively, and 5 days in SOC1 and 6 days in SOC2 groups. The hazard ratio (95% CI) for 2-DG 90 mg group was 3.8 (1.85, 7.94) (*p*=0.0003) compared with the SOC2 group and 3.4 (1.90, 6.01) (*p*<0.0001) compared with the pooled SOC group.

Similarly, time to vital signs normalisation was seen to be significantly better in the 2-DG 90 mg group as compared to SOC2 group. Median time to vital signs normalisation was 5 days in the 2-DG 90 mg group as compared to 8 days in the contemporaneous SOC2 group, Hazard ratio (95% CI) from the CPH model= 4.3 (1.67, 11.29) (*p*=0.0026).

The median time to improvement in clinical status score by 2 points over baseline was 5 days in both 2-DG 90 mg group and 2-DG 126 mg group, even though the median time to improvement by 1 point was shortest in 2-DG 126 mg group (3 days) followed by 2-DG 90 mg group (4 days). The comparisons of these two groups with SOC2 group were not statistically significant. Hazard ratio (95% CI) from CPH model was 1.8 (0.92, 3.36) (*p*=0.0852) for 2-DG 90 mg vs SOC2 group, and 1.18 (0.63, 2.22) (*p*=0.6021) for 2-DG 126 mg vs SOC2 group, for the time to achieve 2 points improvement over baseline in the clinical status score. Only the comparison between the 2-DG 90 mg group and the pooled SOC group in terms of median time to 2 points improvement (HR=2.364; 95% CI: 1.33, 4.18; *p*=0.0032) was found significant.

Median time to first conversion to negative RT-PCR test for SARS-CoV-2 RNA was the shortest in the SOC1 group (3.5 days), followed by the 2-DG 90 mg group with 5.0 days as per CPH model (4.0 days from descriptive statistics) and the 2-DG 63 mg group with 7.0 days as per CPH model (6.0 days from descriptive statistics). The median time was 7.0 days for both the 2-DG 126 mg and SOC2 groups. None of the 2-DG groups was statistically significant compared with the corresponding SOC groups. However, 2-DG 90 mg group showed numerically superior trend with HR= 2.0 (0.94, 4.25; *p*=0.0702).

One patient each in 2-DG 90 mg group and the SOC2 group required ICU admission during the study. One mortality was reported in 2-DG 90 mg group. No meaningful comparison on ICU admission or mortality rates among treatment groups could be made due to the negligible number of events.

### Safety

All three dose levels of 2-DG were well tolerated. A total of 65 treatment-emergent adverse events were reported in 33 (30.3%) patients. One serious adverse event was reported in a patient in the 2-DG 90 mg group who died of ARDS, which was considered not related to the study drug by the sponsor as well as the investigator. Fifty-six of the 65 adverse events (86%) were mild. Hyperglycaemia was the most commonly reported adverse event overall, with 14 events occurring in 10 patients (9.2%) across five groups (Table 3). This included 4 (18.2%) patients in the SOC 2 group, 2 patients each in SOC1 group (9.1%) and 2-DG 126 mg group (9.5%), and 1(4.5%) patient each in 2-DG 63 mg group and 2-DG 90 mg group. Other common adverse events were palpitations in 4 (3.7%) patients, dizziness in 4 (3.7%) patients, and diarrhoea in 3 (2.8%) patients out of 109 patients, all of which were observed in the 2-DG 63 mg and 90 mg groups with incidence ranging 4.5% to 9.1%.

**Table 3:**
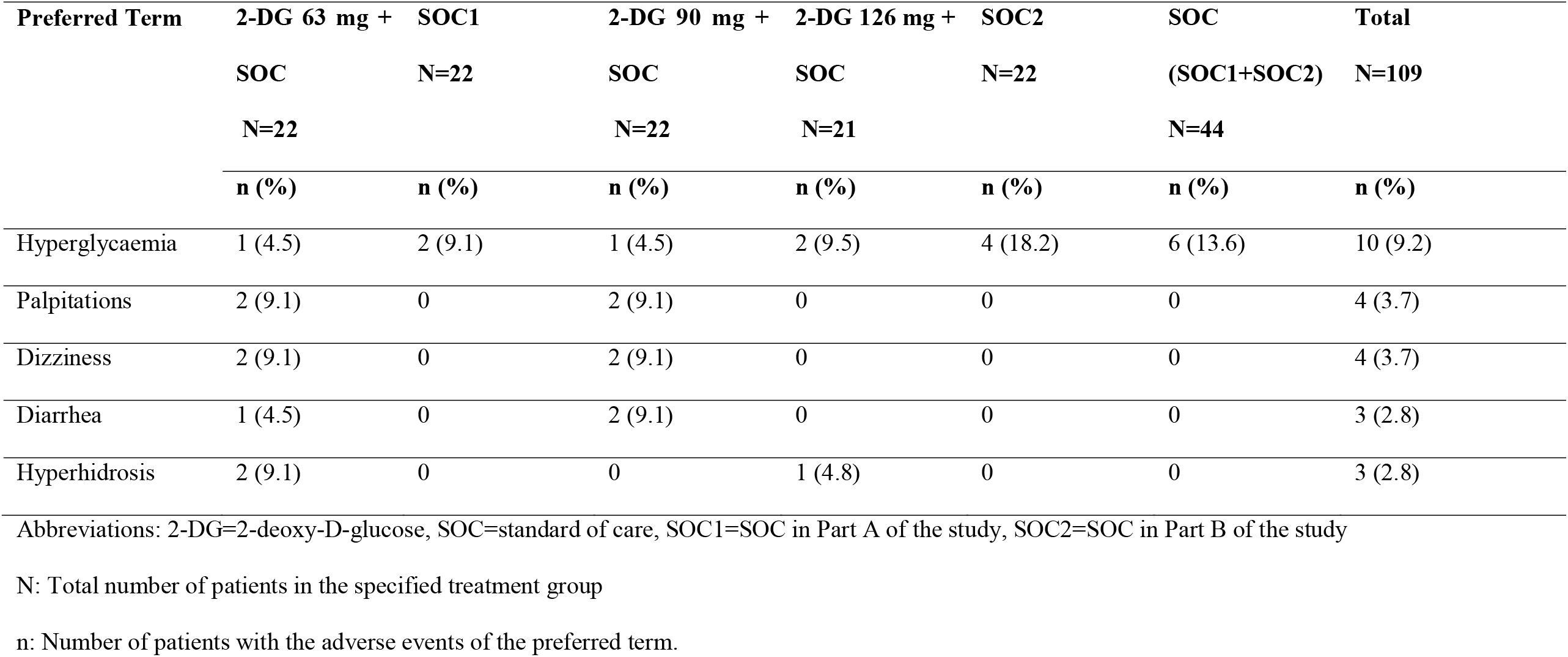
Incidence of treatment-emergent adverse events occurring in >2% of patients.

No clinically significant prolongations of the cardiac QT interval were reported in 2-DG treatment arm. The greatest change in mean QT_c_ intervals from baseline and the highest mean and median values were observed on Day 7 in the 2-DG 126 mg group, with a mean increase of 23.8 msec from baseline (data on file), mean value of 446.7 ms, and median value of 444.0 ms (Table 4).

**Table 4:** Summary of QT_c_B (milliseconds) during the study period.

|  |  | **2-DG 63 mg + SOC** | **SOC1** | **2-DG 90 mg + SOC** | **2-DG 126 mg + SOC** | **SOC2** |
| --- | --- | --- | --- | --- | --- | --- |
| Baseline | N | 22 | 21 | 22 | 21 | 18 |
|  | Mean (SD) | 407.0 (32.79) | 412.5 (29.45) | 429.0 (27.15) | 428.6 (21.21) | 418.9 (33.06) |
|  | Median | 409.5 | 413.0 | 430.0 | 423.0 | 431.5 |
| Day 3 | N | 22 | 22 | 20 | 21 | 21 |
|  | Mean (SD) | 419.1 (24.46) | 412.5 (24.19) | 425.7 (32.72) | 427.5 (29.36) | 427.0 (22.31) |
|  | Median | 413.0 | 406.5 | 423.0 | 427.0 | 421.0 |
| Day 7 | N | 19 | 19 | 10 | 15 | 12 |
|  | Mean (SD) | 415.7 (21.86) | 412.0 (31.39) | 433.5 (30.25) | 446.7 (61.33) | 411.8 (28.40) |
|  | Median | 422.0 | 408.0 | 423.0 | 444.0 | 411.5 |
| End of Treatment | N | 22 | 22 | 19 | 21 | 19 |
|  | Mean (SD) | 413.1 (24.69) | 410.4 (20.61) | 433.9 (37.82) | 435.2 (36.59) | 422.4 (23.20) |
|  | Median | 417.0 | 411.0 | 423.0 | 440.0 | 422.0 |

Abbreviations: 2-DG=2-deoxy-D-glucose, SOC=standard of care, SOC1=SOC in Part A of the study, SOC2=SOC in Part B of the study

## Discussion

Experience suggests that no single agent is sufficient to treat all COVID-19 cases, and a multimodal approach is imperative. In this context, we evaluated 2-DG as a potential therapeutic option for COVID-19 as an adjunct to standard of care. To the best of our knowledge, this is the first clinical study of 2-DG in COVID-19 patients. This study was based on extensive safety evidence for 2-DG from prior clinical studies, anti-viral efficacy of topically applied 2-DG in a herpes simplex study, and in vitro inhibition of SARS-CoV-2 by 2-DG [2-4, 9, 13-15].

The starting dose used in this study, 63 mg/kg/day, was about 4 times lower than the maximum tolerated dose of 250 mg/kg reported in a previous trial of glioblastoma multiforme patients [15]. As COVID-19 management has evolved rapidly, some differences in SOC medications were seen between Parts A and B of the study; however, the SOC was comparable between each active treatment group and the corresponding SOC group during each part of the study.

On several key efficacy endpoints, the most favourable outcomes were observed in the 2-DG 90 mg group as compared with SOC. Statistically significant outcomes were seen with 2-DG 90 mg group in times to achieving and maintaining blood oxygen saturation ≥ 94%, clinical recovery, vital signs normalization and discharge as compared to respective SOC. Few favourable numerical trends were also seen in the 2-DG 126 mg group with regards to achieving blood oxygen saturation ≥ 94%, 1 point and 2 points improvement on the WHO 10-point score, and clinical recovery. However, these were not statistically significant when compared with either contemporaneous SOC or the pooled SOC. Although the pharmacological plausibility of drugs being less effective at higher doses exists, the exact reason cannot be concluded in this case given the small sample size of the study.

The time to achieving and maintaining SpO_2_ ≥ 94% is a clinically meaningful endpoint for COVID-19 drug development [18]. Therefore, the results of this study have established proof-of-concept for 2-DG and justify the evaluation of the 90 mg/kg/day dose of 2-DG in a pivotal phase 3 study. These results also have important implications for the management of moderate to severe COVID-19 patients in the current pandemic context. In an earlier observational study of hospitalised COVID-19 patients in regular hospital ward (i.e., outside of ICU), the average duration on supplemental oxygen was 8 days [19]. In another observational study of severe COVID-19 patients, the median time to getting off supplemental oxygen was 6 days [20]. These data are comparable to what we observed in patients in the SOC groups in our current study, with median time to achieving and maintaining SpO_2_ ≥94% on room air of 5 days. Importantly, the time to SpO2 ≥ 94% on room air was significantly shorter in patients treated with 2-DG 90 mg/kg/day, with median time of 2.5 days (50% reduction). During the massive second wave of COVID-19 in India, shortages in hospital beds with medical-grade oxygen have been experienced at numerous locations. If any therapeutic can help in substantial reduction in supplemental oxygen requirement and hospital bed occupancy in COVID-19 patients, it can go a long way to ease the burden on a country’s healthcare resources.

The postulated biochemical mechanism of action of 2-DG can explain its efficacy in achieving and maintaining blood oxygen saturation. SARS-CoV-2 infects the respiratory tract epithelial cells, which leads to inflammation and impedes the transfer of oxygen in the lungs. As these infected cells have high metabolic demand, 2-DG may accumulate within the infected cells, leading to energy starvation and dearth of anabolic intermediates, which could potentially lead to inhibition of viral replication and host inflammatory response, ultimately translating to clinically meaningful benefits, such as improvement in oxygenation and early recovery. Also, this underlying biochemical mechanism of action would be agnostic to SARS-CoV-2 variants as 2-DG acts on host cell metabolism and does not target the fast-mutating viral proteins [2].

For the 90 mg/kg/day dose of 2-DG, benefits were observed for other efficacy endpoints, including time to discharge from isolation ward, time to clinical recovery, time to vital signs normalisation.

All three dose levels studied were well tolerated and were found to be reasonably safe. The overall incidence of adverse events was low, and the majority of adverse events were mild in intensity. One patient died of ARDS, which was considered not related to 2-DG treatment. Changes in blood glucose levels were evaluated in this study. As observed in previous oncology studies, glycolytic inhibition and competition between glucose and 2-DG for cellular uptake can result in transient hyperglycaemia at higher doses [13]. In this study, the incidence of hyperglycaemia was comparable between the active 2-DG and SOC groups. Moreover, these events of hyperglycaemia were mild and did not lead to study treatment discontinuation in any patients. There was no confirmed adverse event of hypoglycaemia in our study. Maximum plasma concentration achieved with a 45 mg/kg dose of 2-DG was found to be approximately 0.5 mM in an earlier study [14], which is only one-ninth of the plasma glucose concentration (80 mg/dL or 4.5 mM under fasting conditions). Therefore, theoretically the availability of glucose to normal cells, particularly glucose-hungry brain cells, is 9 times higher than 2-DG. It is noteworthy that due to limited mitochondrial function, SARS-CoV-2 infected cells use glycolysis to meet the high bioenergetic and anabolic demand, unlike the uninfected cells that use both glycolysis and mitochondrial respiration to fulfil normal cellular energy demands. 2-DG primarily inhibits glycolysis and its effect on ATP generation from mitochondrial oxidation in normal uninfected cells (without Warburg shift) would be negligible.

Another important safety consideration with 2-DG was its potential for cardiac QT prolongation. While QT prolongation have been reported in previous oncology studies, mostly at higher doses of 2-DG, these effects were transient and asymptomatic [13,14]. In this study, the changes from baseline for mean and median QT_c_ values for 2-DG arm were within acceptable ranges.

A limitation of this phase 2 study is that it was not adequately powered. A subsequent adequately powered (80%) phase 3 clinical study has been initiated with prespecified primary and secondary endpoints. The results from the phase 3 study are expected to be published in the near future.

For several patients in the current phase 2 study, a clinical status score of 4 (‘hospitalised, no oxygen therapy’) was recorded on the WHO 10-point ordinal scale at baseline, despite their requiring oxygen treatment. 21 out of 22 patients had SpO_2_ < 95% at baseline. Due to shortage of oxygen supply, many eligible patients did not receive oxygen supplementation in their respective hospitals. It is possible that these patients were assigned a score of 4 Another limitation was that meaningful comparison between the 2-DG groups and control groups on ICU admission and death was not possible due to small number of such events. In addition, the study was not placebo controlled or blinded.

## Conclusion

Results of the current phase II study suggest 2-DG holds promise as adjunctive treatment to standard of care, in the management of moderate to severe COVID-19 and have encouraged confirmatory evaluation of its efficacy and safety in a larger phase-III clinical trial. If confirmed, 2-DG may provide healthcare practitioners with another option to treat moderate to severe COVID-19 patients.

## Data Availability

Good amount of the data produced in the present work are contained in the manuscript. However, the data which could not be accommodated in the manuscript can be made available upon reasonable request to the authors

## Acknowledgements

The authors wish to thank Dr DM Rao and his team from Dr Reddy’s Laboratories Ltd. for his contributions to all regulatory activities related to 2 DG. The study was conducted by Navitas Life Sciences. The authors would also like to thank all the members of the project team without whom the successful completion of the project would not have been possible.

