## Supplementary for "2-Deoxy-D-Glucose as an Adjunct to Standard of Care in the Medical Management of COVID-19: A Proof-of-Concept & Dose-Ranging Randomised Clinical Trial"

**Supplementary Figure and Tables**

| **Supplementary Figures** |  |
| --- | --- |
| Supplementary Figure 1 | Chemical structures of glucose (left) and 2-deoxy-D-glucose (2-DG) (right) |
| Supplementary Figure 2 | Probability of achieving and maintaining blood oxygen saturation of ≥94% in patients treated with 2-DG 90 mg/kg/day + SOC compared with the contemporaneous SoC group (SOC2) |
| **Supplementary Tables:** |  |
| Supplementary Table 1 | Patient Demographics |
| Supplementary Table 2 | Medical History |
| Supplementary Table 3 | Summary of Time to Clinical Recovery |
| Supplementary Table 4 | Analysis of Time to Clinical Recovery |
| Supplementary Table 5 | Proportion of Patients Showing Negative Conversion of Detectable SARS-CoV-2 RNA on RT-PCR by Study Days 3, 7, 10, and EOT |
| Supplementary Table 6 | Summary of Time to First Negative Conversion of Detectable SARS-CoV-2 By RT-PCR Assay of a Nasopharyngeal/Oropharyngeal Swab Specimen |
| Supplementary Table 7 | Analysis of Time to First Negative Conversion of Detectable SARS-CoV-2 by RT-PCR Assay of a Nasopharyngeal/Oropharyngeal Swab Specimen |
| Supplementary Table 8 | Summary of Time to Discharge from Isolation Ward |
| Supplementary Table 9 | Analysis of Time to Discharge from Isolation Ward |
| Supplementary Table 10 | Summary of Time to Achieve Improvement by At Least 1 and At Least 2 Points Over Baseline in the Clinical Status Score on WHO 10-Point Ordinal Scale from Start of Study Treatment |
| Supplementary Table 11 | Analysis of Median Time (no. of days) to Achieve Improvement by 1 Point and by 2 Points Over Baseline in the Clinical Status Score on WHO 10-Point Ordinal Scale from Start of Study Treatment - Study part A |
| Supplementary Table 12 | Analysis of Median Time (no. of days) to Achieve Improvement by 1 Point and by 2 Points Over Baseline in the Clinical Status Score on WHO 10-Point Ordinal Scale from Start of Study Treatment - Study part B |
| Supplementary Table 13 | Summary of Patients Requiring Management in Intensive Care Unit (ICU), Oxygen Supplementation, Non-Invasive and Invasive Mechanical Ventilation until Day 28 |
| Supplementary Table 14 | Summary of Time to Achieve and Maintain Blood Oxygen Saturation of ≥94% |
| Supplementary Table 15 | Analysis of Time to Achieve and Maintain Blood Oxygen Saturation of ≥94% |
| Supplementary Table 16 | Summary of Time to Vital Signs Normalization |
| Supplementary Table 17 | Analysis of Time to Vital Signs Normalization |
| Supplementary Table 18 | Efficacy endpoint comparisons between active (2-DG) and Pooled SOC groups |
| Supplementary Table 19 | Number and Percentage of patients with TEAEs classified by SOC and PT |

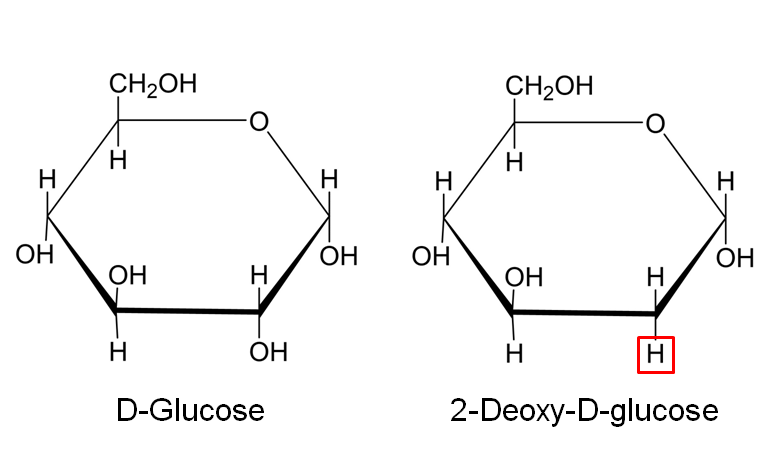

**Supplementary Figure 1: Chemical structures of glucose and 2-deoxy-D-glucose (2-DG)**

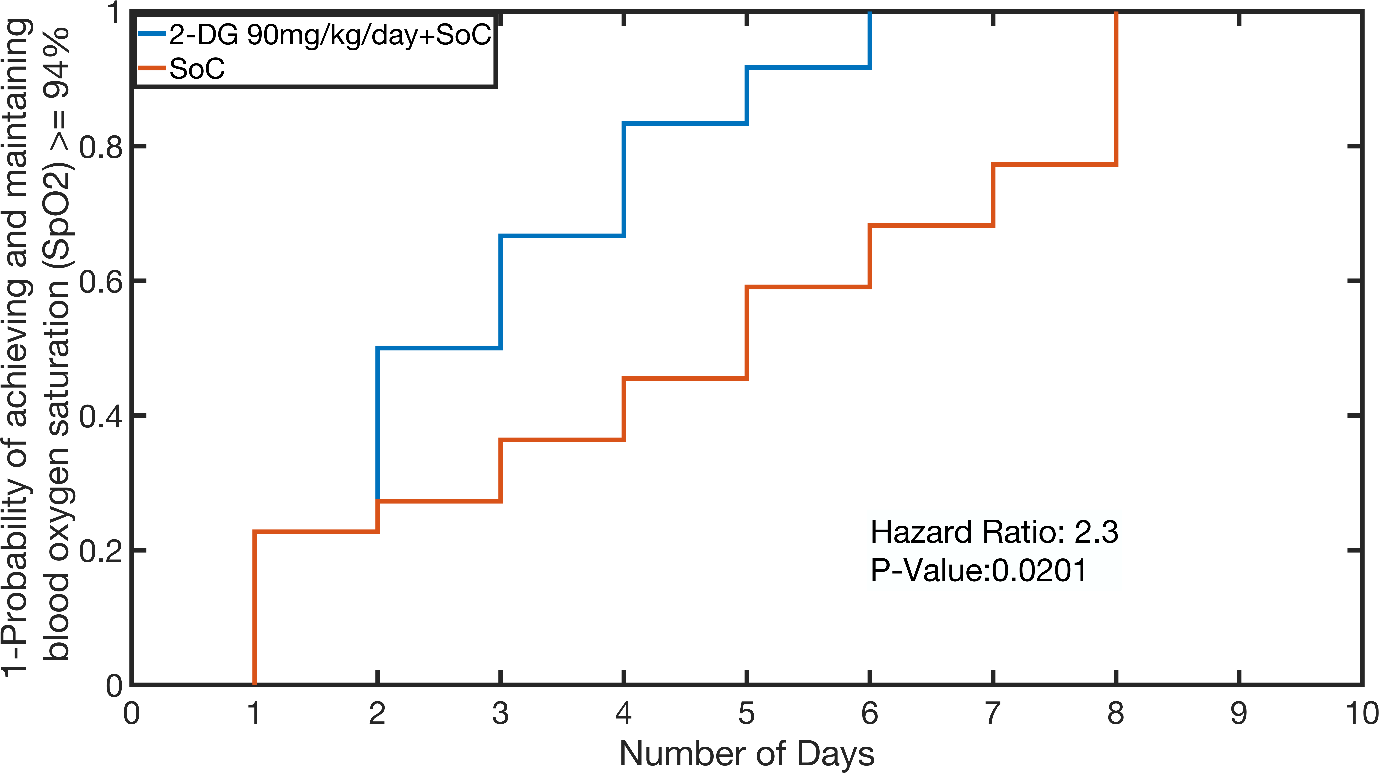

**Supplementary Figure 2: Probability of achieving and maintaining blood oxygen saturation of ≥94% in patients treated with 2-DG 90 mg/kg/day + SOC compared with the contemporaneous SoC group (SOC2)**

Abbreviations: 2-DG=2-deoxy-D-glucose, SOC=standard of care, SOC2=SOC in Part B of the study

**Supplementary Table 1: Patient Demographics**

| **Variable** | **Statistics** | **2-DG 63mg + SOC N=22 n (%)** | **SOC1 N=22 n (%)** | **2-DG 90 mg + SOC N=22 n (%)** | **2-DG 126 mg + SOC N=21 n (%)** | **SOC2  N=22  n (%)** | **Total  N=109  n (%)** |
| --- | --- | --- | --- | --- | --- | --- | --- |
| Age (years) | Mean (SD) | 44.2 (12.71) | 44.4 (9.59) | 46.3 (11.00) | 42.7 (9.33) | 46.6 (11.96) | 44.9 (10.90) |
|  | Median | 45.5 | 42.5 | 45.5 | 41.0 | 47.0 | 44.0 |
| Height (cm) | Mean (SD) | 160.8 (7.38) | 161.0 (7.84) | 166.5 (6.13) | 163.8 (7.76) | 165.7 (8.56) | 163.6 (7.79) |
|  | Median | 161.5 | 161.0 | 167.5 | 162.8 | 164.5 | 164.0 |
| Weight (kg) | Mean (SD) | 61.3 (9.35) | 67.9 (9.76) | 69.4 (10.52) | 74.2 (12.23) | 70.6 (11.77) | 68.6 (11.39) |
|  | Median | 60.3 | 66.0 | 72.5 | 71.3 | 70.3 | 69.0 |
| Sex | Female | 7 (31.8) | 6 (27.3) | 1 (4.5) | 5 (23.8) | 2 (9.1) | 21 (19.3) |
|  | Male | 15 (68.2) | 16 (72.7) | 21 (95.5) | 16 (76.2) | 20 (90.9) | 88 (80.7) |
|  | Transgender | 0 | 0 | 0 | 0 | 0 | 0 |
| Race | Asian | 22 (100.0) | 22 (100.0) | 22 (100.0) | 21 (100.0) | 22 (100.0) | 109 (100.0) |
|  | Others | 0 | 0 | 0 | 0 | 0 | 0 |
| Symptom severity sum score at baseline | n | 22 | 21 | 22 | 21 | 21 | 107 |
|  | Mean (SD) | 4.2 (2.63) | 3.3 (1.83) | 5.4 (3.89) | 6.4 (4.42) | 6.2 (2.93) | 5.1 (3.42) |
|  | Median | 4.0 | 3.0 | 5.5 | 5.0 | 6.0 | 5.0 |

Total column includes all three 2-DG treatment groups, SOC1 and SOC2

N: Total number of patients in the specified treatment group (safety population); n: Total number of patients in the specified treatment group for a given variable

Percentages are based on the total number of patients (safety population).

Symptom severity sum score is calculated for each patient by adding the self-assessed symptom severity score (on 5-Point Likert-type scale) for each COVID-19 associated symptoms (cough, fever, nasal congestion, fatigue, body aches, sore throat, and diarrhoea)

**Supplementary Table 2: Medical History**

| **System Organ Class  Preferred Term** | **2-DG 63 mg+ SOC N=22  n (%)** | **SOC1  N=22  n (%)** | **2-DG 90mg + SOC  N=22 n (%)** | **2-DG 126mg + SOC  N=21 n (%)** | **SOC2  N=22  n (%)** | **Total  N=109  n (%)** |
| --- | --- | --- | --- | --- | --- | --- |
| **Number of patients with medical history** | 10 (45.5) | 13 (59.1) | 2 (9.1) | 4 (19.0) | 2 (9.1) | 31 (28.4) |
| Endocrine disorders | 1 (4.5) | 0 | 0 | 1 (4.8) | 0 | 2 (1.8) |
| Hypothyroidism | 1 (4.5) | 0 | 0 | 1 (4.8) | 0 | 2 (1.8) |
| General disorders and administration site conditions | 1 (4.5) | 1 (4.5) | 0 | 0 | 0 | 2 (1.8) |
| Pyrexia | 1 (4.5) | 1 (4.5) | 0 | 0 | 0 | 2 (1.8) |
| Infections and infestations | 7 (31.8) | 11 (50.0) | 0 | 1 (4.8) | 0 | 19 (17.4) |
| COVID-19 pneumonia | 7 (31.8) | 11 (50.0) | 0 | 1 (4.8) | 0 | 19 (17.4) |
| Investigations | 0 | 0 | 2 (9.1) | 2 (9.5) | 0 | 4 (3.7) |
| Blood creatine phosphokinase increased | 0 | 0 | 1 (4.5) | 0 | 0 | 1 (0.9) |
| Blood lactate dehydrogenase increased | 0 | 0 | 2 (9.1) | 2 (9.5) | 0 | 4 (3.7) |
| C-reactive protein increased | 0 | 0 | 1 (4.5) | 2 (9.5) | 0 | 3 (2.8) |
| White blood cell count increased | 0 | 0 | 0 | 1 (4.8) | 0 | 1 (0.9) |
| Respiratory, thoracic, and mediastinal disorders | 1 (4.5) | 0 | 1 (4.5) | 1 (4.8) | 2 (9.1) | 5 (4.6) |
| Oropharyngeal pain | 1 (4.5) | 0 | 0 | 0 | 0 | 1 (0.9) |
| Rales | 1 (4.5) | 0 | 1 (4.5) | 1 (4.8) | 2 (9.1) | 5 (4.6) |
| Vascular disorders | 2 (9.1) | 2 (9.1) | 0 | 0 | 0 | 4 (3.7) |
| Hypertension | 2 (9.1) | 2 (9.1) | 0 | 0 | 0 | 4 (3.7) |

Total column includes all three 2-DG treatment groups, SOC1 and SOC2

N: Total number of patients in the specified treatment group (safety population)

n: Total number of patients in the specified treatment group for a given category

Patients are counted only once within each SOC class and preferred term

Percentages are based on the total number of patients in the specified treatment group under safety population

**Supplementary Table 3: Summary of Time to Clinical Recovery**

|  | **Statistics** | **2-DG 63 mg + SOC  (N=22)** | **SOC1  (N=22)** | **2-DG 90 mg + SOC  (N=22)** | **2-DG 126 mg + SOC  (N=21)** | **SOC2  (N=22)** | **Pooled SOC (SOC1 + SOC2)  (N=44)** | **Total  (N=109)** |
| --- | --- | --- | --- | --- | --- | --- | --- | --- |
| Time to clinical recovery (Days) | n | 22 | 22 | 19 | 21 | 19 | 41 | 103 |
|  | Mean ± SD | 4.9± 2.76 | 5.4± 2.66 | 3.1± 2.16 | 4.6± 2.79 | 5.8± 3.14 | 5.6± 2.86 | 4.8± 2.81 |
|  | Q1 | 4.0 | 4.0 | 2.0 | 3.0 | 4.0 | 4.0 | 3.0 |
|  | Median | 4.5 | 5.0 | 3.0 | 4.0 | 5.0 | 5.0 | 4.0 |
|  | Q3 | 6.0 | 7.0 | 4.0 | 6.0 | 8.0 | 7.0 | 6.0 |
|  | IQR | 2.0 | 3.0 | 2.0 | 3.0 | 4.0 | 3.0 | 3.0 |
|  | (Min, Max) | (1.0, 13.0) | (1.0, 12.0) | (1.0, 9.0) | (1.0, 10.0) | (1.0, 15.0) | (1.0, 15.0) | (1.0, 15.0) |

Total column includes all three 2-DG treatment groups, SOC1 and SOC2

IQR (interquartile range) = Q3 – Q1

N: Total number of patients in the specified treatment group

n: Total number of patients from each group whose data were used in the calculation of the descriptive summary statistics

Time to Clinical Recovery is a composite endpoint of number of days to achieving and maintaining blood oxygen saturation of ≥94% on room air and the number of days to achieving symptom score of ≤1 (on a 5-point Likert-type scale)for all COVID-19 associated symptoms.

Time to ‘Clinical recovery’= {Date of blood oxygen saturation ≥94% in two consecutive assessments on room air - Date of first dose intake in case of 2-DG/ Date Randomization in case of SOC } +1 **and** Time to ‘Clinical recovery’= {Date of severity score <=1 for all COVID-19 associated symptoms - Date of first dose intake in case of 2-DG/ Date Randomization in case of SOC} +1

COVID-19 associated symptoms whose severity was assessed included cough, fever, nasal congestion, fatigue, body aches, sore throat, and diarrhoea; time to clinical recovery was defined as the earliest time when all symptom severity score first became ‘mild’ (symptom score=1) or ‘absent’ (symptom score=0).

**Supplementary Table 4: Analysis of Time to Clinical Recovery**

|  |  | **2-DG 63 mg + SOC (N=22)** | **SOC1 (N=22)** | **2-DG 90 mg + SOC (N=22)** | **2-DG 126 mg + SOC (N=21)** | **SOC2 (N=22)** |
| --- | --- | --- | --- | --- | --- | --- |
| Time to clinical recovery (Days) | **n** | 22 | 22 | 22 | 21 | 22 |
|  | **Median** | 4.5 | 5 | 3 | 4 | 6 |
|  | **Event n (%)** | 22 (100%) | 22 (100%) | 19 (86%) | 21 (100%) | 19 (86%) |
|  | **Censor n (%)** | 0 | 0 | 3 (14%) | 0 | 3 (14%) |
|  | **SE** | 0.32 |  | 0.371 | 0.364 |  |
|  | **HR (95% CI)** | 0.985 (0.526, 1.846) |  | 3.837 (1.853, 7.944) | 1.881 (0.922, 3.838) |  |
|  | **p-value** | 0.9629 |  | 0.0003 | 0.0824 |  |

CI: Confidence Interval; HR: Hazard Ratio (from Cox Proportional Hazard model) N/n: Total number of patients; SE = Standard Error.

Time to Clinical Recovery is a composite endpoint of number of days to achieving and maintaining blood oxygen saturation of ≥94% on room air and the number of days to achieving symptom score of ≤1 (on a 5-point Likert-type scale)for all COVID-19 associated symptoms.

Time to ‘Clinical recovery’= {Date of blood oxygen saturation ≥94% in two consecutive assessments on room air - Date of first dose intake for 2-DG/ Date of Randomization for SOC } +1 **and** Time to ‘Clinical recovery’= {Date of severity score <=1 for all COVID-19 associated symptoms - Date of first dose intake for 2-DG/ Date of Randomization for SOC} +1

HR a its corresponding 95% CI and two-sided p-value were obtained using the Cox proportional hazard model with age, gender, and baseline symptom score as covariate.

HR, its corresponding 95% CI, and two-sided p-value were obtained by comparing 2-DG 63 mg with SOC1 and comparing 2-DG 90 mg and 126 mg with SOC2.

Patients who did not achieve clinical recovery from baseline up to Day 28 were right-censored.

**Supplementary Table 5: Proportion of Patients Showing Negative Conversion of Detectable SARS-CoV-2 RNA on RT-PCR by Study Days 3, 7, 10, and EOT**

|  | | **2-DG 63mg + SOC  N=22** | | | **SOC1  N=22** | | | **2-DG 90mg + SOC  N=22** | | | **2-DG 126mg + SOC  N=21** | | | **SOC2  N=22** | | | **SOC (SOC1+SOC2)  N=44** | | | **Total  N=109** | |
| --- | --- | --- | --- | --- | --- | --- | --- | --- | --- | --- | --- | --- | --- | --- | --- | --- | --- | --- | --- | --- | --- |
| Timepoint | Patients completed visit | | n (%) | Patients completed visit | | n (%) | Patients completed visit | | n (%) | Patients completed visit | | n (%) | Patients completed visit | | n (%) | Patients completed visit | | n (%) | Patients completed visit | | n (%) |
| Day 3 | 22 | | 9 (40.9) | 22 | | 12 (54.5) | 20 | | 6 (27.3) | 21 | | 6 (28.6) | 21 | | 3 (13.6) | 43 | | 15 (34.1) | 106 | | 36 (33.0) |
| Day 7 | 18 | | 17 (77.3) | 19 | | 16 (72.7) | 10 | | 10 (45.5) | 15 | | 8 (38.1) | 12 | | 7 (31.8) | 31 | | 23 (52.3) | 74 | | 58 (53.2) |
| Day 10 | 12 | | 18 (81.8) | 12 | | 21 (95.5) | 8 | | 14 (63.6) | 13 | | 16 (76.2) | 8 | | 9 (40.9) | 20 | | 30 (68.2) | 53 | | 78 (71.6) |
| EOT | 22 | | 20 (90.9) | 22 | | 22 (100.0) | 22 | | 15 (68.2) | 21 | | 18 (85.7) | 22 | | 13 (59.1) | 44 | | 35 (79.5) | 109 | | 88 (80.7) |

N: Total number of patients in the specified treatment group

n: Total number of patients with negative conversion (of detectable SARS-CoV-2 viral RNA) on nasopharyngeal/oropharyngeal swab.

Total column includes all three 2-DG treatment groups, SOC1 and SOC2

**Supplementary Table 6: Summary of Time to First Negative Conversion of detectable SARS-CoV-2 by RT-PCR assay of a nasopharyngeal/oropharyngeal swab specimen**

|  | **Statistics** | **2-DG 63 mg + SOC  (N=22)** | **SOC1  (N=22)** | **2-DG 90 mg + SOC  (N=22)** | **2-DG 126 mg + SOC  (N=21)** | **SOC2  (N=22)** | **SOC (SOC1 + SOC2)  (N=44)** | **Total  (N=109)** |
| --- | --- | --- | --- | --- | --- | --- | --- | --- |
| Time to first negative conversion (Days) | n | 20 | 22 | 20 | 21 | 19 | 41 | 102 |
|  | Mean ± SD | 5.7± 2.94 | 6.0± 3.76 | 5.0± 3.06 | 7.0± 4.01 | 6.9± 3.31 | 6.4± 3.54 | 6.1± 3.46 |
|  | Q1 | 3.0 | 3.0 | 3.0 | 3.0 | 4.0 | 3.0 | 3.0 |
|  | Median | 6.0 | 3.5 | 4.0 | 7.0 | 7.0 | 7.0 | 6.0 |
|  | Q3 | 7.0 | 10.0 | 7.0 | 10.0 | 9.0 | 9.0 | 9.0 |
|  | IQR | 4.0 | 7.0 | 4.0 | 7.0 | 5.0 | 6.0 | 6.0 |
|  | (Min, Max) | (3.0, 12.0) | (3.0, 15.0) | (1.0, 10.0) | (1.0, 14.0) | (1.0, 15.0) | (1.0, 15.0) | (1.0, 15.0) |

Total column includes all three 2-DG treatment groups, SOC1 and SOC2

IQR= Q3 – Q1

N: Total number of patients in the specified treatment group

n: Total number of patients from each group whose data were used in the calculation of the descriptive summary statistics

Time to negative conversion = {Date of negative conversion on nasopharyngeal/oropharyngeal swab – Date of first dose intake in case of 2-DG/ Date of randomization in case of SOC} +1

**Supplementary Table 7: Analysis of Time to First Negative Conversion of detectable SARS-CoV-2 by RT-PCR assay of a nasopharyngeal/oropharyngeal swab specimen**

|  |  | **2-DG 63 mg + SOC (N=22)** | **SOC1 (N=22)** | **2-DG 90 mg + SOC (N=22)** | **2-DG 126 mg + SOC (N=21)** | **SOC2 (N=22)** |
| --- | --- | --- | --- | --- | --- | --- |
| Time to first negative conversion (Days) | **n** | 22 | 22 | 22 | 21 | 22 |
|  | **Median** | 7 | 3.5 | 5 | 7 | 7 |
|  | **Event n (%)** | 20 (91%) | 22 (100%) | 20 (91%) | 21 (100%) | 19 (86%) |
|  | **Censor, n (%)** | 2 (9%) | 0 | 2 (9%) | 0 | 3 (14%) |
|  | **SE** | 0.329 |  | 0.384 | 0.344 |  |
|  | **HR (95% CI)** | 0.926 (0.486, 1.765) |  | 2.003 (0.944, 4.251) | 1.354 (0.69, 2.656) |  |
|  | **p-value** | 0.8158 |  | 0.0702 | 0.3787 |  |

CI: Confidence Interval; HR: Hazard Ratio (from Cox Proportional Hazard model) N/n: Total number of patients; SE: Standard Error

Time to negative conversion is the time from start of study treatment to time of attaining the negative conversion (of detectable SARS CoV-2 viral RNA) on swab (nasopharyngeal/ oropharyngeal)

Time to negative conversion = {Date of first negative conversion on nasopharyngeal/oropharyngeal swab – Date of first dose intake for 2-DG/Date of Randomization date for SOC} +1

HR, its corresponding 95% CI and two-sided p-value were obtained using the Cox proportional hazard model with viral load (RdRp Target) at baseline as covariate.

HR, its corresponding 95% CI, and two-sided p-value were obtained by comparing 2-DG 63 mg with SOC1 and 2-DG 90 mg and 126mg with SOC2.

Patients who had not achieved SARS-CoV-2 negative from baseline up to day 28 were right-censored.

**Supplementary Table 8: Summary of Time to Discharge from Isolation Ward**

|  | **Statistics** | **2-DG 63 mg + SOC  (N=22)** | **SOC1  (N=22)** | **2-DG 90 mg + SOC  (N=22)** | **2-DG 126 mg + SOC  (N=21)** | **SOC2  (N=22)** | **SOC (SOC1 + SOC2)  (N=44)** | **Total  (N=109)** |
| --- | --- | --- | --- | --- | --- | --- | --- | --- |
| Time to discharge from isolation ward (Days) | n | 19 | 22 | 19 | 20 | 20 | 42 | 100 |
|  | Mean ± SD | 11.3± 3.56 | 10.6± 2.90 | 7.9± 3.27 | 10.7± 5.22 | 9.4± 3.72 | 10.0± 3.34 | 10.0± 3.91 |
|  | Q1 | 9.0 | 9.0 | 5.0 | 7.0 | 7.0 | 8.0 | 7.0 |
|  | Median | 11.0 | 11.0 | 8.0 | 11.0 | 9.0 | 10.0 | 11.0 |
|  | Q3 | 13.0 | 12.0 | 11.0 | 13.0 | 12.0 | 12.0 | 12.0 |
|  | IQR | 4.0 | 3.0 | 6.0 | 6.0 | 5.0 | 4.0 | 5.0 |
|  | (Min, Max) | (5.0, 21.0) | (6.0, 16.0) | (3.0, 12.0) | (4.0, 27.0) | (3.0, 18.0) | (3.0, 18.0) | (3.0, 27.0) |

Total column includes all three treatment groups, SOC1 and SOC2

IQR = Q3 – Q1

N: Total number of patients in the specified treatment group

n: Total number of patients from each group whose data were used in the calculation of the descriptive summary statistics

Time to discharge from isolation ward is the time from start of study treatment intake to discharge from the isolation ward.

Time to discharge from isolation ward = {Date of discharge from the isolation ward – Date of first dose intake for 2-DG/ Date of Randomization date for SOC} +1

**Supplementary Table 9: Analysis of Time to Discharge from Isolation Ward**

|  |  | **2-DG 63 mg + SOC (N=22)** | **SOC1 (N=22)** | **2-DG 90 mg + SOC (N=22)** | **2-DG 126 mg + SOC (N=21)** | **SOC2 (N=22)** |
| --- | --- | --- | --- | --- | --- | --- |
| Time to Discharge from Isolation Ward (Days) | **n** | 22 | 22 | 22 | 21 | 22 |
|  | **Median** | 12 | 11 | 8 | 11 | 10 |
|  | **Event n (%)** | 19 (86%) | 22(100%) | 19 (86%) | 20 (95%) | 20 (91%) |
|  | **Censor n (%)** | 3 (14%) | 0 | 3 (14%) | 1 (5%) | 2 (9%) |
|  | **SE** | 0.328 |  | 0.379 | 0.362 |  |
|  | **HR (95% CI)** | 0.791 (0.416, 1.504) |  | 2.238 (1.065, 4.703) | 0.679 (0.334, 1.38) |  |
|  | **p-value** | 0.4746 |  | 0.0336 | 0.2847 |  |

CI: Confidence Interval; HR: Hazard Ratio (from Cox Proportional Hazard model) N/n: Total number of patients; SE: Standard Error

Time to discharge from isolation ward is the time from start of study treatment to discharge from the isolation ward.

Time to discharge from isolation ward = {Date of discharge from the isolation ward – Date of first dose intake for 2-DG/ Date of Randomization date for SOC} +1

HR, its corresponding 95% CI and two-sided p-value were obtained using the Cox proportional hazard model with age, gender, and viral load (RdRp Target) at baseline as covariate

HR, its corresponding 95% CI, and two-sided p-value were obtained by comparing 2-DG 63 mg with SOC1 and 2-DG 90 mg and 2-DG 126 mg with SOC2.

Patients who had not achieved discharge from the isolation ward from baseline up to day 28 were right-censored.

**Supplementary Table 10 - Summary of Time to Achieve Improvement by At Least 1 Point and At Least 2 Points over Baseline in the Clinical Status Score on WHO 10-Point Ordinal Scale** **from Start of Study Treatment**

| **Time to improvement in clinical status** | **Statistics** | **2-DG 63 mg + SOC  (N=22)** | **SOC1  (N=22)** | **2-DG 90 mg + SOC  (N=22)** | **2-DG 126 mg + SOC  (N=21)** | **SOC2  (N=22)** | **SOC (SOC1 + SOC2)  (N=44)** | **Total  (N=109)** |
| --- | --- | --- | --- | --- | --- | --- | --- | --- |
| Improvement by 1 point (Days) | n | 20 | 22 | 19 | 21 | 20 | 42 | 102 |
|  | Mean ± SD | 5.3± 2.08 | 6.4± 2.59 | 4.0± 1.91 | 4.6± 3.53 | 4.8± 2.12 | 5.6± 2.49 | 5.0± 2.62 |
|  | Q1 | 4.0 | 4.0 | 2.0 | 2.0 | 3.0 | 4.0 | 3.0 |
|  | Median | 5.0 | 6.0 | 4.0 | 3.0 | 5.0 | 5.0 | 5.0 |
|  | Q3 | 6.5 | 8.0 | 6.0 | 5.0 | 6.0 | 7.0 | 6.0 |
|  | IQR | 2.5 | 4.0 | 4.0 | 3.0 | 3.0 | 3.0 | 3.0 |
|  | (Min, Max) | (2.0, 10.0) | (2.0, 11.0) | (1.0, 8.0) | (1.0, 14.0) | (2.0, 9.0) | (2.0, 11.0) | (1.0, 14.0) |
| Improvement by 2 points (Days) | n | 19 | 21 | 19 | 21 | 20 | 41 | 100 |
|  | Mean ± SD | 11.0± 3.90 | 10.1± 2.56 | 5.0± 2.11 | 6.0± 3.94 | 6.3± 2.12 | 8.2± 3.05 | 7.7± 3.84 |
|  | Q1 | 9.0 | 9.0 | 3.0 | 4.0 | 5.0 | 6.0 | 5.0 |
|  | Median | 11.0 | 10.0 | 5.0 | 5.0 | 6.0 | 8.0 | 7.0 |
|  | Q3 | 13.0 | 11.0 | 7.0 | 7.0 | 8.0 | 10.0 | 10.0 |
|  | IQR | 4.0 | 2.0 | 4.0 | 3.0 | 3.0 | 4.0 | 5.0 |
|  | (Min, Max) | (4.0, 21.0) | (6.0, 15.0) | (2.0, 9.0) | (1.0, 18.0) | (2.0, 10.0) | (2.0, 15.0) | (1.0, 21.0) |

Total column includes all three 2-DG treatment groups, SOC1 and SOC2

IQR= Q3 – Q1

N: Total number of patients in the specified treatment group

n: Total number of patients from each group whose data were used in the calculation of the descriptive summary statistics

Time to improvement in clinical status score by at least 1-point/2-points uses WHO 10-point ordinal scale to assess the improvement from start of the study treatment.

Time to improvement in clinical status score by at least 1-point after randomization = {Date of at least 1-point drop in Clinical Status score on WHO 10-point ordinal scale - Date of first dose intake for 2-DG/ Date of Randomization date for SOC}+1

Time to improvement in clinical status score by at least 2-points after randomization = {Date of at least 2-point drop in Clinical Status score on WHO 10-point ordinal scale e - Date of first dose intake for 2-DG/ Date of Randomization date for SOC}+1

**Supplementary Table 11: Analysis of Median Time to Achieving Improvement by 1 point and by 2 Points over Baseline in the Clinical Status Score on WHO 10-Point Ordinal Scale from Start of Study Treatment - Study Part A**

|  | | **n** | **Median** | **Event n (%)** | **Censor n (%)** | **SE** | **HR (95% CI)** | **p-value** |
| --- | --- | --- | --- | --- | --- | --- | --- | --- |
| Time to 1-point improvement (Days) | 2-DG 63 mg + SOC (N=22) | 22 | 5.5 | 20 (91%) | 2 (9%) | 0.352 | 1.414 (0.71, 2.818) | 0.3247 |
|  | SOC1 (N=22) | 22 | 6 | 22 (100%) | 0 |  |  |  |
|  | Baseline WHO 10-point ordinal scale |  |  |  |  | 0.449 | 0.895 (0.371, 2.159) | 0.8048 |
| Time to 2-point improvement (Days) | 2-DG 63 mg + SOC(N=22) | 22 | 11 | 19 (86%) | 3 (14%) | 0.344 | 0.624 (0.318, 1.224) | 0.1702 |
|  | SOC1(N=22) | 22 | 10.5 | 21 (95%) | 1 (5%) |  |  |  |
|  | Baseline WHO 10-point ordinal scale |  |  |  |  | 0.677 | 4.751 (1.26, 17.912) | 0.0214 |

CI: Confidence Interval; HR: Hazard Ratio (from Cox Proportional Hazard model) N n: Total number of patients; SE: Standard Error

Time to improvement in clinical status score by at least 1-point/2-points uses WHO 10-point ordinal scale to assess the improvement from start of the study treatment.

Time to improvement in clinical status score by at least 1-point = {Date of at least 1-point drop in Clinical Status score on WHO 10-point ordinal scale - Date of first dose intake for 2-DG/ Date of Randomization date for SOC} +1

Time to improvement in clinical status score by at least 2-points = {Date of at least 2-point drop in Clinical Status score on WHO 10-point ordinal scale e - Date of first dose intake for 2-DG/ Date of Randomization date for SOC}+1

HR, its corresponding 95% CI and two-sided p-value were obtained using the Cox proportional hazard model with WHO 10-point ordinal scale clinical

status score at baseline as covariate.

HR, its corresponding 95% CI, and two-sided p-value were obtained by comparing 2-DG 63 mg with SOC1 and 2-DG 90 mg and 120 mg with SOC2.

Patients whose clinical status score had not improved by 1 point and by 2 points from baseline up to day 28 were right-censored.

**Supplementary Tab****le 12: Analysis of Median Time to Achieving Improvement by 1 point and by 2 points over baseline in the Clinical Status Score on WHO 10-Point Ordinal Scale from Start of Study Treatment - Study Part B**

|  | | **n** | **Median** | **Event n (%)** | **Censor n (%)** | **SE** | **HR (95% CI)** | **p-value** |
| --- | --- | --- | --- | --- | --- | --- | --- | --- |
| Time to 1-point improvement (Days) | 2-DG 90 mg + SOC (N=22) | 22 | 4 | 19 (86%) | 3 (14%) | 0.328 | 1.483 (0.78, 2.822) | 0.2297 |
|  | 2-DG 126 mg + SOC (N=21) | 21 | 3 | 21 (100%) | 0 | 0.318 | 1.301 (0.697, 2.427) | 0.4090 |
|  | SOC2 (N=22) | 22 | 5 | 20 (91%) | 2 (9%) |  |  |  |
|  | Baseline WHO 10-point ordinal scale |  |  |  |  | 0.288 | 0.812 (0.462, 1.427) | 0.4685 |
| Time to 2-point improvement (Days) | 2-DG 90 mg + SOC (N=22) | 22 | 5 | 19 (86%) | 3 (14%) | 0.329 | 1.763 (0.924, 3.363) | 0.0852 |
|  | 2-DG 126 mg + SOC (N=21) | 21 | 5 | 21 (100%) | 0 | 0.322 | 1.183 (0.63, 2.221) | 0.6021 |
|  | SOC2 (N=22) | 22 | 6 | 20 (91%) | 2 (9%) |  |  |  |
|  | Baseline WHO 10-point ordinal scale |  |  |  |  | 0.294 | 0.681 (0.383, 1.212) | 0.1917 |

CI: Confidence Interval; HR: Hazard Ratio (from Cox Proportional Hazard model) N /n: Total number of patients; SE: Standard Error

Time to improvement in clinical status score by at least 1-point/2-points uses WHO 10-point ordinal scale to assess the improvement from start of the study treatment.

Time to improvement in clinical status score by at least 1-point = {Date of at least 1-point drop in Clinical Status score on WHO 10-point ordinal scale - Date of first dose intake for 2-DG/ Date of Randomization date for SOC} +1

Time to improvement in clinical status score by at least 2-points = {Date of at least 2-point drop in Clinical Status score on WHO 10-point ordinal scale e - Date of first dose intake for 2-DG/ Date of Randomization date for SOC}+1

HR, its corresponding 95% CI and two-sided p-value were obtained using the Cox proportional hazard model with WHO 10-point ordinal scale clinical

status score at baseline as covariate.

HR, its corresponding 95% CI, and two-sided p-value were obtained by comparing 2-DG 63 mg with SOC1 and 2-DG 90 mg and 120 mg with SOC2.

Patients whose clinical status score had not improved by 1 point and by 2 points from baseline up to day 28 were right-censored.

**Supplementary Table 13: Summary of Patients Requiring Management in Intensive Care Unit (ICU), Oxygen Supplementation, Non-invasive and Invasive Mechanical Ventilation until End of Treatment**

|  | **2-DG 63mg + SOC  N=22** | | **SOC1  N=22** | | **2-DG 90mg + SOC  N=22** | | **2-DG 126mg + SOC  N=21** | | **SOC2  N=22** | | **SOC (SOC1+SOC2)  N=44** | | **Total  N=109** | |
| --- | --- | --- | --- | --- | --- | --- | --- | --- | --- | --- | --- | --- | --- | --- |
|  | **n (%)** | **m (%)** | **n (%)** | **m (%)** | **n (%)** | **m (%)** | **n (%)** | **m (%)** | **n (%)** | **m (%)** | **n (%)** | **m (%)** | **n (%)** | **m (%)** |
| Patients requiring treatment of Management in intensive care unit (ICU) | 0 | 0 | 0 | 0 | 1 (4.5) | 1 (4.5) | 0 | 0 | 1 (4.5) | 1 (4.5) | 1 (2.3) | 1 (2.3) | 2 (1.8) | 2 (1.8) |
| Patients requiring oxygen supplementation | 20 (90.9) | 0 | 22 (100.0) | 0 | 8 (36.4) | 1 (4.5) | 6 (28.6) | 1 (4.8) | 9 (40.9) | 0 | 31 (70.5) | 0 | 65 (59.6) | 2 (1.8) |
| Patients requiring invasive mechanical ventilation | 0 | 0 | 0 | 0 | 0 | 0 | 1 (4.8) | 0 | 0 | 0 | 0 | 0 | 1 (0.9) | 0 |
| Patients requiring non-invasive mechanical ventilation | 0 | 0 | 0 | 0 | 1 (4.5) | 1 (4.5) | 0 | 0 | 1 (4.5) | 1 (4.5) | 1 (2.3) | 1 (2.3) | 2 (1.8) | 2 (1.8) |

Total column includes all three treatment groups, SOC1 and SOC2

N: Total number of patients in the specified treatment group

n: Total number of patients in the specified category; m: Total number of patients in the specified category after treatment start date

**Supplementary T****able 14: Summary of Time to Achieve and Maintain Blood Oxygen Saturation ≥94%**

|  | **Statistics** | **2-DG 63 mg + SOC  (N=22)** | **SOC1  (N=22)** | **2-DG 90 mg + SOC  (N=22)** | **2-DG 126 mg + SOC  (N=21)** | **SOC2  (N=22)** | **SOC (SOC1 + SOC2)  (N=44)** | **Total  (N=109)** |
| --- | --- | --- | --- | --- | --- | --- | --- | --- |
| Time to achieve and maintain blood oxygen saturation (Days) | n | 22 | 20 | 19 | 21 | 21 | 41 | 103 |
|  | Mean ± SD | 5.3± 3.87 | 5.3± 2.66 | 2.6± 1.46 | 4.6± 3.09 | 4.5± 2.64 | 4.9± 2.65 | 4.5± 3.00 |
|  | Q1 | 4.0 | 4.0 | 1.0 | 3.0 | 2.0 | 3.0 | 2.0 |
|  | Median | 5.0 | 5.0 | 2.0 | 3.0 | 5.0 | 5.0 | 4.0 |
|  | Q3 | 6.0 | 6.0 | 4.0 | 7.0 | 7.0 | 6.0 | 6.0 |
|  | IQR | 2.0 | 2.0 | 3.0 | 4.0 | 5.0 | 3.0 | 4.0 |
|  | (Min, Max) | (1.0, 17.0) | (1.0, 12.0) | (1.0, 6.0) | (1.0, 12.0) | (1.0, 8.0) | (1.0, 12.0) | (1.0, 17.0) |

Total column includes all three 2-DG treatment groups, SOC1 and SOC2

IQR= Q3 – Q1

N: Total number of patient’s in the specified treatment group

n: Total number of patients from each group whose data were used in the calculation of the descriptive summary statistics

Time to achieve and maintain blood oxygen saturation ≥ 94% is the time (days) from randomization/first dose intake to reach the oxygen saturation of at least 94% on room air.

Time to achieve and maintain blood oxygen saturation ≥ 94% = {Date blood oxygen saturation first became ≥94%on two consecutive evaluations - Date of first dose intake for 2-DG/ Date of randomization for SOC} +1

**Supplementary Table 15: Analysis of Time to Achieve and Maintain Blood Oxygen Saturation ≥94%**

|  |  |  | **2-DG 63 mg + SOC (N=22)** | **SOC1 (N=22)** | **2-DG 90 mg + SOC (N=22)** | **2-DG 126 mg + SOC (N=21)** | **SOC2 (N=22)** |
| --- | --- | --- | --- | --- | --- | --- | --- |
|  | Analysis of Time to Maintain Blood Oxygen Saturation (Days) | **n** | 22 | 22 | 22 | 21 | 22 |
|  |  | **Median (Days)** | 5 | 5 | 2.5 | 3 | 5 |
|  |  | **Event n (%)** | 22 (100%) | 20 (91%) | 19 (86%) | 21 (100%) | 21 (95%) |
|  |  | **Censor n (%)** | 0 | 2 (9%) | 3 (14%) | 0 | 2 (9%) |
|  |  | **SE** | 0.338 |  | 0.358 | 0.347 |  |
|  |  | **HR(LL-UL)** | 1.277 (0.658, 2.477) |  | 2.3 (1.14, 4.642) | 0.975 (0.494, 1.925) |  |
|  |  | **p-value** | 0.4698 |  | 0.0201 | 0.9415 |  |
| CI: Confidence Interval; HR: Hazard Ratio (from Cox Proportional Hazard model) N /n: Total number of patients in the ITT population; SE: Standard Error | | | | | | | |
| Time to achieve and maintain blood oxygen saturation ≥ 94% is the time (days) from randomization/first dose intake to reach the oxygen saturation of at least 94% on room air. | | | | | | | |
| Time to achieve and maintain blood oxygen saturation ≥ 94% = {Date blood oxygen saturation first became ≥94%on two consecutive evaluations - Date of first dose intake for 2-DG/ Date of randomization for SOC} +1  HR, its corresponding 95% CI and two-sided p-value were obtained using the Cox proportional hazard model with age, gender, and baseline blood oxygen saturation as covariate. | | | | | | | |
| HR, its corresponding 95% CI, and two-sided p-value were obtained by comparing 2-DG 63 mg with SOC1 and 2-DG 90 mg and 120 mg with SOC2. | | | | | | | |
| Patients who had not achieved blood oxygen saturation ≥94% from baseline up to day 28 were right-censored. | | | | | | | |

**Supplementary Table 16: Summary of Time to Vital Signs Normalization**

|  | **Statistics** | **2-DG 63 mg + SOC  (N=22)** | **SOC1  (N=22)** | **2-DG 90 mg + SOC  (N=22)** | **2-DG 126 mg + SOC  (N=21)** | **SOC2  (N=22)** | **SOC (SOC1 + SOC2)  (N=44)** | **Total  (N=109)** |
| --- | --- | --- | --- | --- | --- | --- | --- | --- |
| Time to Vital Signs normalization (Days) | N | 20 | 22 | 12 | 11 | 9 | 31 | 74 |
|  | Mean ± SD | 6.7± 2.87 | 6.5± 2.67 | 5.2± 2.66 | 7.8± 3.09 | 6.0± 1.41 | 6.4± 2.36 | 6.5± 2.72 |
|  | Q1 | 5.0 | 4.0 | 3.0 | 6.0 | 5.0 | 5.0 | 5.0 |
|  | Median | 7.0 | 7.0 | 5.0 | 6.0 | 6.0 | 6.0 | 6.0 |
|  | Q3 | 8.5 | 8.0 | 5.0 | 10.0 | 7.0 | 8.0 | 8.0 |
|  | IQR | 3.5 | 4.0 | 2.0 | 4.0 | 2.0 | 3.0 | 3.0 |
|  | (Min, Max) | (1.0, 13.0) | (2.0, 12.0) | (3.0, 11.0) | (4.0, 13.0) | (4.0, 8.0) | (2.0, 12.0) | (1.0, 13.0) |

Total column includes all three 2-DG treatment groups, SOC1 and SOC2

IQR= Q3 – Q1

N: Total number of patient’s in the specified treatment group

n: Total number of patients from each group whose data were used in the calculation of the descriptive summary statistics

Time to vital signs normalization is a composite score when all the following vital signs parameter were satisfied (i.e., Body temperature below 98.9℉, respiratory rate below 20 breaths per minute, blood oxygen saturation (SpO2) >95 percentage on room air and heart rate below 90 bpm after start of study treatment) from start of the study treatment.

Time to Vital Signs normalization = {Earliest date when all following vital sign parameters are satisfied body temperature below 98.9ºF, Respiratory rate below 20 breaths per minute, Blood oxygen saturation (SpO2) >95% on room air and Heart rate below 90 bpm – Date of first dose intake for 2-DG/ Date of Randomization date for SOC} +1

**Supplementary Table 17: Time to Vital Signs Normalization**

|  |  |  | **2-DG 63 mg + SOC (N=22)** | **SOC1 (N=22)** | **2-DG 90 mg + SOC (N=22)** | **2-DG 126 mg + SOC (N=21)** | **SOC2 (N=22)** |
| --- | --- | --- | --- | --- | --- | --- | --- |
|  | Analysis of Time to Vital Signs Normalization (Days) | **n** | 22 | 22 | 22 | 21 | 22 |
|  |  | **Median (Days)** | 7 | 7 | 5 | 10 | 8 |
|  |  | **Event n (%)** | 20 (91%) | 22(100%) | 12 (55%) | 11 (52%) | 9 (41%) |
|  |  | **Censor n (%)** | 2 (9%) | 0 | 10 (45%) | 10 (48%) | 13 (59%) |
|  |  | **SE** | 0.323 |  | 0.488 | 0.458 |  |
|  |  | **HR (95% CI)** | 0.889 (0.472, 1.674) |  | 4.341 (1.669, 11.294) | 1.024 (0.418, 2.511) |  |
|  |  | **p-value** | 0.7162 |  | 0.0026 | 0.9580 |  |
| CI: Confidence Interval; HR: Hazard Ratio (from Cox Proportional Hazard model) N /n: Total number of patient’s in the ITT population; SE: Standard Error | | | | | | | |
| Time to vital signs normalization is a composite score when all the following vital signs parameter were satisfied (i.e., Body temperature below 98.9℉, respiratory rate below 20 breaths per minute, blood oxygen saturation (SpO2) >95 percentage on room air and heart rate below 90 bpm after start of study treatment) from start of the study treatment. | | | | | | | |
| Time to Vital Signs normalization = {Earliest date when all following vital sign parameters are satisfied body temperature below 98.9ºF, Respiratory rate below 20 breaths per minute, Blood oxygen saturation (SpO2) >95% on room air and Heart rate below 90 bpm – Date of first dose intake for 2-DG/ Date of Randomization date for SOC} +1 | | | | | | | |
| HR, its corresponding 95% CI and 2-sided p-value were obtained using the Cox proportional hazard model with Clinical Status score on WHO 10-point | | | | | | | |
| ordinal scale at baseline as covariate. | | | | | | | |
| HR, its corresponding 95% CI, and two-sided p-value were obtained by comparing 2-DG 63 mg with SOC1 and 2-DG 90 mg and 120 mg with SOC2. | | | | | | | |
| Patients who had not achieved Vital Sign normalization up to Day 28 were right-censored. | | | | | | | |

**Supplementary Table 18: Efficacy endpoint comparisons between active (2-DG) and Pooled SOC groups**

|  |  | **2-DG 63 mg + SOC** | **SOC1** | **2-DG 90 mg + SOC** | **2-DG 126 mg + SOC** | **SOC2** | **2-DG 90 +126 mg +SOC** | **SOC Pool (SOC1 + SOC2)** |
| --- | --- | --- | --- | --- | --- | --- | --- | --- |
| Time (days) to Achieve SpO2 ≥94% for two consecutive time on room air | N | 22 | 22 | 22 | 21 | 22 | 43 | 44 |
|  | Median (Days) | 5 | 5 | 2.5 | 3 | 5 | 3 | 5 |
|  | HR (95% CI) ^a^ | 1.01 (0.59, 1.73) |  | 2.651 (1.49, 4.73) | 1.132 (0.66, 1.95) |  | 1.56 (0.99, 2.45) |  |
|  | p-value^a^ | 0.98 |  | 0.0009 | 0.66 |  | 0.057 |  |
| Time (days) to Discharge from Isolation Ward | Median (Days) | 12 | 11 | 8 | 11 | 10 | 11 | 10 |
|  | HR (95% CI) ^a^ | 0.735 (0.42, 1.28) |  | 2.209 (1.21, 4.04) | 0.822 (0.47, 1.44) |  | 1.17 (0.74, 1.86) |  |
|  | p-value^a^ | 0.2748 |  | 0.01 | 0.4934 |  | 0.5049 |  |
| Time (days) to Vital Signs Normalisation^b^ | Median (Days) | 7 | 7 | 5 | 10 | 8 | 9 | 7 |
|  | HR (95% CI) ^a^ | 0.856 (0.47, 1.55) |  | 2.16(1.08, 4.33) | 0.703  (0.35, 1.41) |  | 1.086 (0.63, 1.89) |  |
|  | p-value^a^ | 0.6082 |  | 0.0301 | 0.3214 |  | 0.7709 |  |
| Time (days) to achieve 2 Point improvement on WHO 10-Point Scale | Median (Days) | 11 | 10.5 | 5 | 5 | 6 | 5 | 8 |
|  | HR (95% CI) ^a^ | 0.811 (0.46, 1.44) |  | 2.361  (1.33, 4.18) | 1.543 (0.90, 2.64) |  | 1.85 (1.18, 2.91) |  |
|  | p-value^a^ | 0.4772 |  | 0.0032 | 0.114 |  | 0.0077 |  |
| Time (days) to achieve 1 Point improvement on WHO 10-Point Scale | Median (Days) | 5.5 | 6 | 4 | 3 | 5 | 4 | 5 |
|  | HR (95% CI) ^a^ | 1.289  (0.74, 2.26) |  | 1.63  (0.93, 2.85) | 1.284  (0.75, 2.19) |  | 1.429  (0.92, 2.23) |  |
|  | p-value^a^ | 0.3765 |  | 0.087 | 0.3605 |  | 0.1155 |  |

Abbreviations: CI=confidence interval, 2-DG=2-deoxy-D-glucose, HR=hazard ratio, SOC=standard of care, SOC1=SOC in Part A of the study, SOC2=SOC in Part B of the study, WHO=World Health Organization

^a^Each 2-DG+SOC group was compared with pooled SOC (SOC1+SOC2), in Parts A and B of the study.

^b^Time to vital signs normalisation was defined as the earliest date when all the following vital signs parameters were satisfied: body temperature <98.9℉, respiratory rate <20 breaths per minute, blood oxygen saturation (SpO2) >95% on room air and heart rate <90 bpm after start of study treatment.

**Supplementary Table 19: Number and Percentage of Patients with TEAEs Classified by System Organ Class and Preferred Term**

|  | **2-DG 63mg + SOC  N=22** | | **SOC1  N=22** | | **2-DG 90mg + SOC  N=22** | | **2-DG 126mg + SOC  N=21** | | **SOC2  N=22** | | **SOC (SOC1+SOC2)  N=44** | | **Total  N=109** | |
| --- | --- | --- | --- | --- | --- | --- | --- | --- | --- | --- | --- | --- | --- | --- |
| **System Organ Class**  **Preferred Term** | **Number of Events** | **n (%)** | **Number of Events** | **n (%)** | **Number of Events** | **n (%)** | **Number of Events** | **n (%)** | **Number of Events** | **n (%)** | **Number of Events** | **n (%)** | **Number of Events** | **n (%)** |
| Number of patients with at least one AE | 17 | 9 (40.9) | 11 | 6 (27.3) | 22 | 8 (36.4) | 6 | 5 (23.8) | 9 | 5 (22.7) | 20 | 11 (25.0) | 65 | 33 (30.3) |
| Cardiac disorders | 2 | 2 (9.1) | 0 | 0 | 3 | 3 (13.6) | 1 | 1 (4.8) | 0 | 0 | 0 | 0 | 6 | 6 (5.5) |
| Palpitations | 2 | 2 (9.1) | 0 | 0 | 2 | 2 (9.1) | 0 | 0 | 0 | 0 | 0 | 0 | 4 | 4 (3.7) |
| Sinus tachycardia | 0 | 0 | 0 | 0 | 1 | 1 (4.5) | 1 | 1 (4.8) | 0 | 0 | 0 | 0 | 2 | 2 (1.8) |
| Gastrointestinal disorders | 2 | 2 (9.1) | 1 | 1 (4.5) | 5 | 3 (13.6) | 0 | 0 | 0 | 0 | 1 | 1 (2.3) | 8 | 6 (5.5) |
| Abdominal pain | 0 | 0 | 0 | 0 | 1 | 1 (4.5) | 0 | 0 | 0 | 0 | 0 | 0 | 1 | 1 (0.9) |
| Constipation | 0 | 0 | 1 | 1 (4.5) | 0 | 0 | 0 | 0 | 0 | 0 | 1 | 1 (2.3) | 1 | 1 (0.9) |
| Diarrhoea | 1 | 1 (4.5) | 0 | 0 | 2 | 2 (9.1) | 0 | 0 | 0 | 0 | 0 | 0 | 3 | 3 (2.8) |
| Nausea | 0 | 0 | 0 | 0 | 2 | 2 (9.1) | 0 | 0 | 0 | 0 | 0 | 0 | 2 | 2 (1.8) |
| Vomiting | 1 | 1 (4.5) | 0 | 0 | 0 | 0 | 0 | 0 | 0 | 0 | 0 | 0 | 1 | 1 (0.9) |
| General disorders and administration site conditions | 0 | 0 | 3 | 3 (13.6) | 4 | 2 (9.1) | 0 | 0 | 0 | 0 | 3 | 3 (6.8) | 7 | 5 (4.6) |
| Chills | 0 | 0 | 1 | 1 (4.5) | 0 | 0 | 0 | 0 | 0 | 0 | 1 | 1 (2.3) | 1 | 1 (0.9) |
| Fatigue | 0 | 0 | 0 | 0 | 3 | 2 (9.1) | 0 | 0 | 0 | 0 | 0 | 0 | 3 | 2 (1.8) |
| Pain | 0 | 0 | 1 | 1 (4.5) | 1 | 1 (4.5) | 0 | 0 | 0 | 0 | 1 | 1 (2.3) | 2 | 2 (1.8) |
| Swelling face | 0 | 0 | 1 | 1 (4.5) | 0 | 0 | 0 | 0 | 0 | 0 | 1 | 1 (2.3) | 1 | 1 (0.9) |
| Infections and infestations | 2 | 2 (9.1) | 0 | 0 | 0 | 0 | 0 | 0 | 0 | 0 | 0 | 0 | 2 | 2 (1.8) |
| Viral myocarditis | 2 | 2 (9.1) | 0 | 0 | 0 | 0 | 0 | 0 | 0 | 0 | 0 | 0 | 2 | 2 (1.8) |
| Investigations | 1 | 1 (4.5) | 0 | 0 | 2 | 2 (9.1) | 1 | 1 (4.8) | 0 | 0 | 0 | 0 | 4 | 4 (3.7) |
| Blood glucose increased | 1 | 1 (4.5) | 0 | 0 | 0 | 0 | 0 | 0 | 0 | 0 | 0 | 0 | 1 | 1 (0.9) |
| Electrocardiogram QT prolonged | 0 | 0 | 0 | 0 | 1 | 1 (4.5) | 1 | 1 (4.8) | 0 | 0 | 0 | 0 | 2 | 2 (1.8) |
| Electrocardiogram T wave inversion | 0 | 0 | 0 | 0 | 1 | 1 (4.5) | 0 | 0 | 0 | 0 | 0 | 0 | 1 | 1 (0.9) |
| Metabolism and nutrition disorders | 2 | 1 (4.5) | 5 | 3 (13.6) | 1 | 1 (4.5) | 2 | 2 (9.5) | 5 | 4 (18.2) | 10 | 7 (15.9) | 15 | 11 (10.1) |
| Decreased appetite | 0 | 0 | 1 | 1 (4.5) | 0 | 0 | 0 | 0 | 0 | 0 | 1 | 1 (2.3) | 1 | 1 (0.9) |
| Hyperglycaemia | 2 | 1 (4.5) | 4 | 2 (9.1) | 1 | 1 (4.5) | 2 | 2 (9.5) | 5 | 4 (18.2) | 9 | 6 (13.6) | 14 | 10 (9.2) |
| Musculoskeletal and connective tissue disorders | 1 | 1 (4.5) | 0 | 0 | 0 | 0 | 0 | 0 | 1 | 1 (4.5) | 1 | 1 (2.3) | 2 | 2 (1.8) |
| Pain in extremity | 1 | 1 (4.5) | 0 | 0 | 0 | 0 | 0 | 0 | 1 | 1 (4.5) | 1 | 1 (2.3) | 2 | 2 (1.8) |
| Nervous system disorders | 3 | 2 (9.1) | 1 | 1 (4.5) | 4 | 3 (13.6) | 0 | 0 | 1 | 1 (4.5) | 2 | 2 (4.5) | 9 | 7 (6.4) |
| Ageusia | 0 | 0 | 0 | 0 | 1 | 1 (4.5) | 0 | 0 | 0 | 0 | 0 | 0 | 1 | 1 (0.9) |
| Burning sensation | 1 | 1 (4.5) | 0 | 0 | 0 | 0 | 0 | 0 | 0 | 0 | 0 | 0 | 1 | 1 (0.9) |
| Dizziness | 2 | 2 (9.1) | 0 | 0 | 3 | 2 (9.1) | 0 | 0 | 0 | 0 | 0 | 0 | 5 | 4 (3.7) |
| Headache | 0 | 0 | 1 | 1 (4.5) | 0 | 0 | 0 | 0 | 1 | 1 (4.5) | 2 | 2 (4.5) | 2 | 2 (1.8) |
| Psychiatric disorders | 2 | 1 (4.5) | 1 | 1 (4.5) | 0 | 0 | 0 | 0 | 0 | 0 | 1 | 1 (2.3) | 3 | 2 (1.8) |
| Insomnia | 2 | 1 (4.5) | 0 | 0 | 0 | 0 | 0 | 0 | 0 | 0 | 0 | 0 | 2 | 1 (0.9) |
| Stress | 0 | 0 | 1 | 1 (4.5) | 0 | 0 | 0 | 0 | 0 | 0 | 1 | 1 (2.3) | 1 | 1 (0.9) |
| Respiratory, thoracic, and mediastinal disorders | 0 | 0 | 0 | 0 | 3 | 3 (13.6) | 0 | 0 | 2 | 1 (4.5) | 2 | 1 (2.3) | 5 | 4 (3.7) |
| Acute respiratory distress syndrome | 0 | 0 | 0 | 0 | 1 | 1 (4.5) | 0 | 0 | 0 | 0 | 0 | 0 | 1 | 1 (0.9) |
| Cough | 0 | 0 | 0 | 0 |  |  |  |  |  |  |  |  |  |  |
| Hypoxia | 0 | 0 | 0 | 0 | 0 | 0 | 0 | 0 | 1 | 1(4.5) | 1 | 1 (2.3) | 1 | 1 (0.9) |
| Nasal congestion | 0 | 0 | 0 | 0 | 0 | 0 | 0 | 0 | 1 | 1 (4.5) | 1 | 1 (2.3) | 1 | 1 (0.9) |
| Skin and subcutaneous tissue disorders | 2 | 2 (9.1) | 0 | 0 | 0 | 0 | 2 | 1 (4.8) | 0 | 0 | 0 | 0 | 4 | 3 (2.8) |
| Hyperhidrosis | 2 | 2 (9.1) | 0 | 0 | 0 | 0 | 2 | 1 (4.8) | 0 | 0 | 0 | 0 | 4 | 3 (2.8) |

N = Total number of patients in the specified treatment group (safety population); n: number of patients with at least one TEAE in the specified field

Subjects are counted only once within each Preferred Term and System Organ Class

Percentages are based on the total number of patients in the specified treatment group within the Safety Population

Adverse events were coded using the Medical Dictionary of Regulatory Activities (MedDRA version 23.0).
